# Assessing the effectiveness of near real-time flow cytometry in monitoring ozone disinfection in a full-scale drinking water treatment plant

**DOI:** 10.1101/2023.12.29.23300640

**Authors:** Katherine S. Dowdell, Kirk Olsen, Ernesto F. Martinez Paz, Aini Sun, Jeff Keown, Rebecca Lahr, Brian Steglitz, Andrea Busch, John J. LiPuma, Terese Olson, Lutgarde Raskin

**Affiliations:** Department of Civil and Environmental Engineering, University of Michigan, 1353 Beal Ave., Ann Arbor, MI 48109, USA; Ann Arbor Water Treatment Plant, City of Ann Arbor, 919 Sunset Rd., Ann Arbor, MI 48103, USA; Great Lakes Water Authority, 9300 W. Jefferson Ave, Detroit, MI 48209, USA; Department of Pediatrics, University of Michigan Medical School, 8323 MSRB III, SPC5646, 1150 W. Med Cntr Dr., Ann Arbor, MI 48109, USA

**Keywords:** Flow cytometry, Disinfection, Real-time microbial monitoring, Drinking water, Ozone

## Abstract

While real-time monitoring of physicochemical parameters has widely been incorporated into drinking water treatment systems, real-time microbial monitoring has lagged behind, resulting in the use of surrogate parameters (disinfectant residual, applied dose, concentration × time [CT]) to assess disinfection system performance. Near real-time flow cytometry (NRT-FCM) allows for automated quantification of total and intact microbial cells but has not been widely implemented in full-scale systems. This study sought to investigate the feasibility of NRT-FCM for full-scale drinking water ozone disinfection system performance monitoring. A water treatment plant with high lime solids turbidity in the ozone contactor influent was selected to evaluate the NRT-FCM in challenging conditions. Total and intact cell counts were monitored for 40 days and compared to surrogate parameters (ozone residual, ozone dose, and CT) and grab sample assay results for cellular adenosine triphosphate (cATP), heterotrophic plate counts (HPC), impedance flow cytometry, and 16S rRNA gene sequencing. NRT-FCM provided insight into the dynamics of the full-scale ozone system, including offering early warning of increased contactor effluent cell concentrations, which was not observed using surrogate measures. A strong correlation between log intact cell removal and CT was also not observed (Kendall’s tau= −0.09, p=0.04). Positive correlations were observed between intact cell counts and cATP levels (Kendall’s tau=0.40, p<0.01), HPC (Kendall’s tau=0.20, p<0.01), and impedance flow cytometry results (Kendall’s tau=0.30, p<0.01). However, 16S rRNA gene sequencing results showed that passage through the ozone contactor significantly changed the microbial community (p<0.05), supporting the hypothesis that regrowth was occurring in the later chambers of the contactor. This study demonstrates the utility of direct, near real-time microbial analysis for monitoring full-scale disinfection systems.

**Graphical Abstract:** 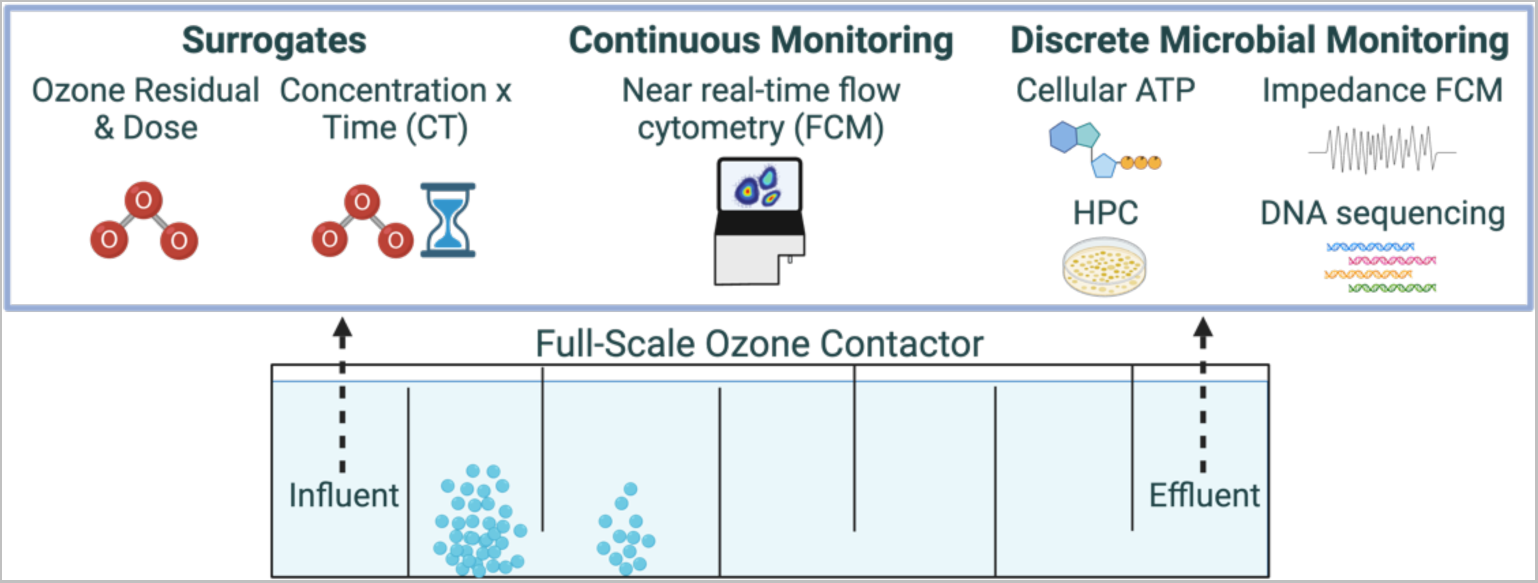

**Highlights:**

- Near real-time flow cytometry (NRT-FCM) was effective for ozone system monitoring.
- Intact microbial cell counts were consistent with cellular ATP and HPC results.
- NRT-FCM provided early detection of increased effluent cell concentrations.
- The ozone contactor influent and effluent microbial communities were distinct.

## 1. Introduction

Real-time monitoring of drinking water treatment processes is crucial to producing high-quality water and protecting public health. However, despite the wide adoption of real-time monitoring for physicochemical parameters in drinking water treatment, automated monitoring of microbiological parameters in full-scale systems has lagged behind (Banna et al., 2014; Storey et al., 2011). Reasons for the slow adoption of real-time or near real-time microbial monitoring include technological and operational challenges and regulations that require culture-based methods (Safford and Bischel, 2019). Microbial monitoring is particularly important for drinking water disinfection systems, where changes in influent composition or process performance upsets can impact the microbiological quality of finished water (Khan et al., 2015; Risebro et al., 2007; Storey et al., 2011). Utilities typically rely on a combination of culture-based methods and real-time monitoring of physicochemical surrogates (e.g., disinfectant residual) to monitor disinfection system performance. However, culture-based methods are slow and only detect a small portion of the microorganisms present in a sample (Allen et al., 2004; Hammes et al., 2008; Van Nevel et al., 2017), and surrogate monitoring may be impacted by water quality and reactor design (Morrison et al., 2022).

Cultivation-independent methods for drinking water microbial monitoring include measures of overall microbial activity (e.g., enzyme activity, cellular adenosine triphosphate [cATP] concentration), DNA-based methods, and direct cell counting methods (e.g., microscopy, flow cytometry [FCM]). Methods that monitor enzyme activity have targeted a variety of enzymes, typically measuring fluorescence as a proxy for total enzyme activity (Burnet et al., 2019; Keon et al., 2021; Lautenschlager et al., 2014). cATP, a universal energy molecule in cells, is quantified using the luciferase enzyme and luminometers (de Vera and Wert, 2019; Magic-Knezev and van der Kooij, 2004). DNA-based methods employed in drinking water include quantitative polymerase chain reaction and DNA sequencing, with the bacterial 16S rRNA gene used as the most common target (Kirisits et al., 2019; Zhang and Liu, 2019). FCM uses microfluidics to pass individual cells through the path of a laser, providing microbial cell counts and information about cell characteristics such as size and shape. In traditional drinking water applications of FCM, cells are stained with fluorescent dyes that bind with nucleic acids (Berney et al., 2008; Hoefel et al., 2003; Phe et al., 2005). Impedance FCM, which quantifies intact cells by passing them through an electric field, offers the potential of intact cell counting without fluorescent staining, but has not been tested in drinking water applications (Cheung et al., 2010; Clausen et al., 2018). FCM, which is primarily used in drinking water to determine total cell counts (TCC) or intact cell counts (ICC), is an attractive option to monitor disinfection processes because it directly quantifies cells. In contrast, enzyme activity, cATP, and DNA-based methods rely on conversion factors to estimate cell concentration (Van Nevel et al., 2017). Additionally, FCM provides data on particle characteristics and may be used to make inferences about cell viability and the microbial community (Favere et al., 2020). Of the microbial monitoring methods, cATP, enzyme activity, and FCM have been automated and deployed at the full-scale level (Besmer and Hammes, 2016; Buysschaert et al., 2018; de Vera and Wert, 2019; Favere et al., 2020; Prest et al., 2021; Sylvestre et al., 2021).

Near real-time FCM (NRT-FCM) includes the automation of sample collection and staining, enabling near real-time microbial monitoring. NRT-FCM has been previously tested in pilot-scale and full-scale drinking water systems for monitoring source waters, membrane filtration systems, chlorine contact tanks, and distribution systems (Buysschaert et al., 2018; Cheswick et al., 2022; Gabrielli et al., 2021; Props et al., 2018), but has not been utilized for full-scale ozone disinfection system monitoring. However, FCM monitoring of ozone inactivation has been evaluated at the bench-scale level (Ramseier et al., 2011), reporting rapid decreases in total and intact cells, even at low ozone doses. Ozone reacts quickly with cellular components, damaging nucleic acids and cell membranes (Hunt and Mariñas, 1999, 1997; von Sonntag and von Gunten, 2012). Therefore, processes like NRT-FCM that detect changes to membrane integrity may be ideal for monitoring ozone disinfection system performance.

In the United States, drinking water ozone disinfection system performance is regulated using the ozone residual concentration, which is then used with system flow properties (flow rate, pH, and temperature) to calculate the log removal of target pathogens, such as *Cryptosporidium* spp., viruses, and *Giardia* spp. (US EPA, 1999). The log removal is determined using concentration × time (CT), which quantifies overall disinfectant exposure. While there are several methods for calculating CT, one of the most common is the T10 method, in which the time required for 10% of the contactor water volume to pass through the system is calculated (US EPA, 2010a). The determination of CT relies on both previously established inactivation relationships and flow dynamics of ozone contactors. However, the numerous factors that impact disinfection system performance, such as the physicochemical properties of the water and non-ideal flow within reactors, can be challenging to incorporate into disinfection calculations (Elovitz et al., 2000; Morrison et al., 2022; Zhang et al., 2007). Additionally, changes in system flow, placement of ozone sensors or sampling lines, and maintenance of ozone sensors have also been shown to impact ozone residual concentrations, which then influence calculated CT values (von Gunten, 2003; Zhang et al., 2007). Therefore, there is a need for methods that facilitate the rapid, direct characterization of full-scale ozone system performance.

This study sought to evaluate NRT-FCM for direct monitoring of full-scale drinking water ozonation. The experiment was conducted for 40 days at a facility previously reported to have unique challenges with the ozone contactors due to lime softening solids carryover (Kotlarz et al., 2018). Thus, it was an ideal location to implement near real-time microbial monitoring. In addition to NRT-FCM, real-time sensors were employed to monitor physicochemical parameters (Martinez Paz et al., 2022). This study also compared the results of NRT-FCM to established and novel methods of measuring system performance and changes in cell viability and activity, including cATP, HPC, impedance FCM, and 16S rRNA gene sequencing. The ozone system performance, characterized using direct microbial methods, was compared to the performance quantified using surrogate parameters (ozone residual, CT).

## 2. Material and methods

### 2.1 Experimental overview and drinking water treatment plant

The NRT-FCM system testing was conducted for 40 days, from late April 2021 to early June 2021. Follow-up testing, which consisted of HPC analysis, occurred during the summer of 2022. Testing was conducted at the City of Ann Arbor Water Treatment Plant, a 50 million gallon per day (MGD, 1.9×10^5^ m^3^/d) facility in Ann Arbor, MI, USA, that serves approximately 125,000 customers. The plant has been described previously (Kotlarz et al., 2018; Pinto et al., 2012). Briefly, groundwater is mixed with Huron River water (approximately 15% groundwater) and treated upstream of the ozone system using two-stage excess lime softening, coagulation, flocculation, and sedimentation. Lime is added to the primary softening basins, and a coagulant, polydiallyldimethylammonium chloride (PolyDADMAC), is added to the secondary softening basins. The water pH is then decreased to approximately 7.5 using carbon dioxide gas before entering the ozone system. After ozonation, the water is filtered using granular activated carbon [GAC] biological filters, disinfected with monochloramine, and fluorinated prior to distribution. The plant recently installed an intermittently operated ultraviolet (UV) system to meet log reduction requirements for *Cryptosporidium* spp. when operated with single-stage lime softening during maintenance periods to meet the US EPA’s Long-Term 2 Surface Water Treatment Rule disinfection requirements. Though the plant has a capacity of 50 MGD, water production during the study ranged from 9 to 23 MGD (3.4×10^4^ to 8.7×10^4^ m^3^/d).

During the 2021 testing period, several operational changes occurred at the water treatment plant, including substantial flow rate changes (Figure S1). Due to maintenance requirements from solids accumulation in the lime softening basins, the plant switches from one set of basins to the other in late spring or early summer, a process that results temporarily in elevated turbidity in the lime softening effluent. The switch of lime softening basins occurred on April 20, 2021, one week before the start of the study. This switch resulted in higher ozone contactor influent turbidity and alkalinity. The median alkalinity during the week before the basin switch was 54 mg CaCO3/L, while the median alkalinity the week after the switch was 86 mg CaCO_3_/L. A PolyDADMAC feed is also started each spring to prevent zebra mussel growth in the river water intake. PolyDADMAC feeding began during the experiment on May 20, 2021 (Day 23), a day at which higher than usual ozone contactor influent turbidity was also observed. No other significant process changes, other than those that were part of the study, occurred during the testing period.

### 2.2 Ozone system, compliance, and CT determination

The ozone system consists of four parallel ozone contactors, of which two are typically in service at a time. Each contactor consists of seven cells (Cell 1 - 7), separated by vertical baffles (Figure 1). All cells are 1,200 ft^3^ (34 m^3^) except for Cell 3, which is 2,400 ft^3^ (68 m^3^). Ozone is applied in Cells 2 and 3 using ceramic fine bubble diffusers with ozone gas generated on-site. Typically, 70% of the ozone gas is sent to Cell 2 in a counter-flow regime (in the opposite direction of water flow), and the remaining 30% is sent to Cell 3 in a co-flow regime (ozone gas and water flow in the same direction). The ozone system is operated to achieve a 0.5 log removal credit for *Giardia* spp. Ozone dosing is controlled by adjusting the ozone content in the feed gas and the gas flow rate. The target ozone residual in Cell 2 of the contactor is at least 0.3 mg/L. Grab samples are collected for compliance monitoring every four hours from the effluents of Cells 2 to 5 for each contactor in operation, and an online ozone residual probe is used to periodically monitor Cells 2, 3, and 4. The ozone system CT is calculated using the grab sample ozone residual measurements in Cells 2, 3, and 4 and the effective contact time (T10), as described in the US EPA Long-Term 2 Enhanced Surface Water Treatment Rule (Equations S1 – S4)(US EPA, 2010a). The applied ozone dose was calculated using the percent ozone in the gas feed, the gas flow rate, and the plant flow rate (Equation S5). The ozone transfer efficiency used by the plant in calculations is 95%. Additional details of ozone calculations are provided in the supplementary information.

**Figure 1.**
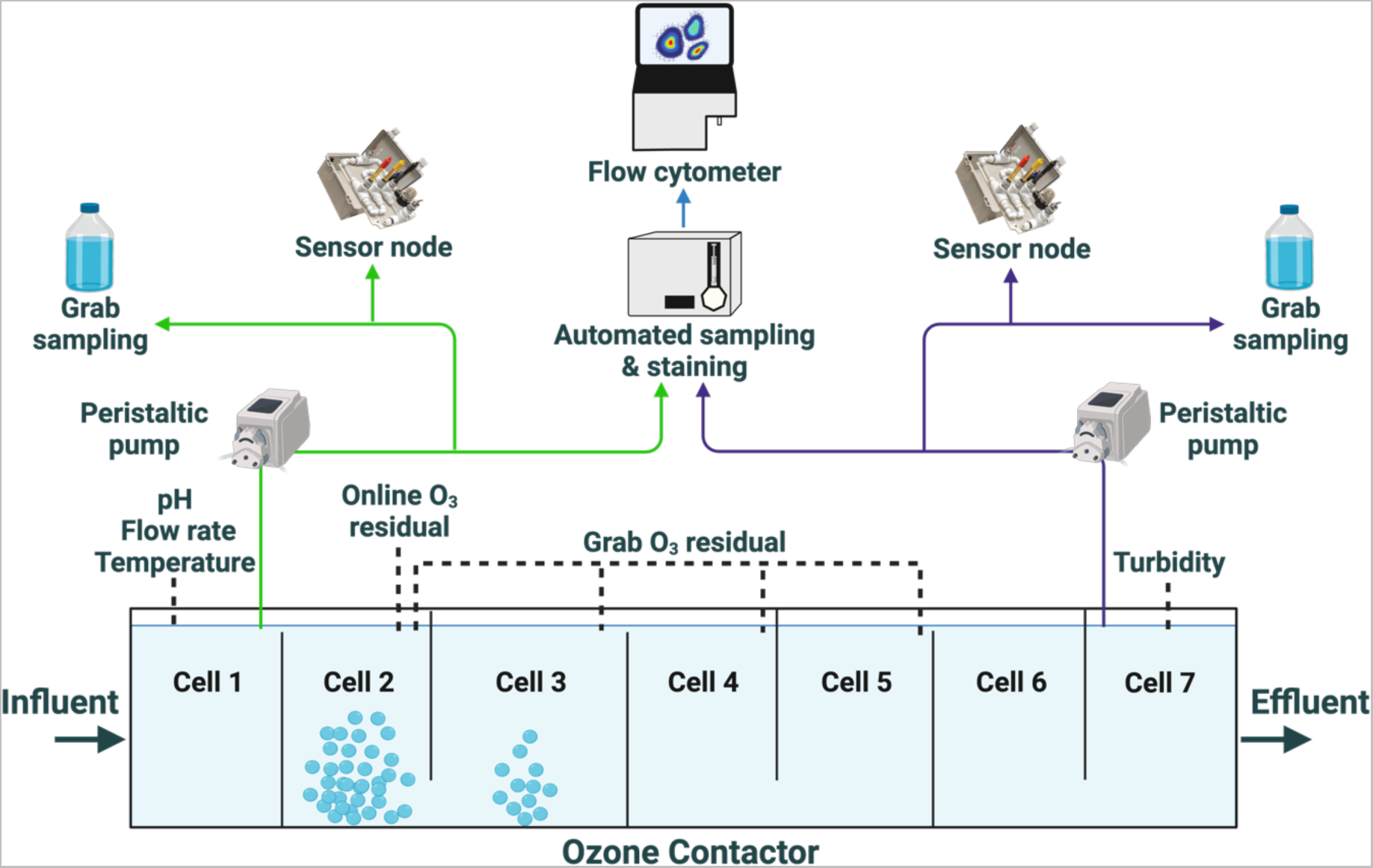
Online monitoring instrumentation set-up for the experiment. Existing monitoring by the water treatment plant that was utilized included real-time monitoring of the influent pH, flow rate, and temperature, online ozone residual monitoring, ozone residual grab samples from Cells 2, 3, and 4, and effluent turbidity. The near real-time flow cytometry system used for the experiment collected and analyzed samples from the influent and effluent; the sensor nodes added for this study monitored pH, temperature, oxidation-reduction potential, and electrical conductivity in the influent and effluent in real time. Prepared using BioRender.com.

Two of the four contactors were in service during the study (Contactors 1 and 3), and each contactor was treating approximately 50% of the total plant flow. All testing for this study was performed on Contactor 1, which had been offline for approximately five months before its start-up in April 2021. Due to lime solids carryover and other particulate matter in the ozone contactor influent, the ozone contactors are periodically taken out of service and cleaned. Contactor 1 was last cleaned in summer 2020, or approximately one year before testing. After the NRT-FCM study, the contactor was taken out of service. It was repaired and cleaned from November to December 2021 and placed back into service. The follow-up sampling occurred during summer 2022.

### 2.3 Ozone challenge testing

To investigate the performance of the NRT-FCM system at different ozone residuals, the target ozone residual at the effluent of Cell 2 was increased for two weeks, beginning on the morning of Day 28. The operators were asked to maintain a Cell 2 ozone residual of at least 0.6 mg/L O_3_ through Day 31 (Ozone Challenge Test 1), then asked to maintain a residual of at least 0.8 mg/L O_3_ from Days 31 through 38 (Ozone Challenge Test 2; Table 1). The target ozone residual was reduced to the normal level of 0.3 mg/L O_3_ on Day 38.

**Table 1.**
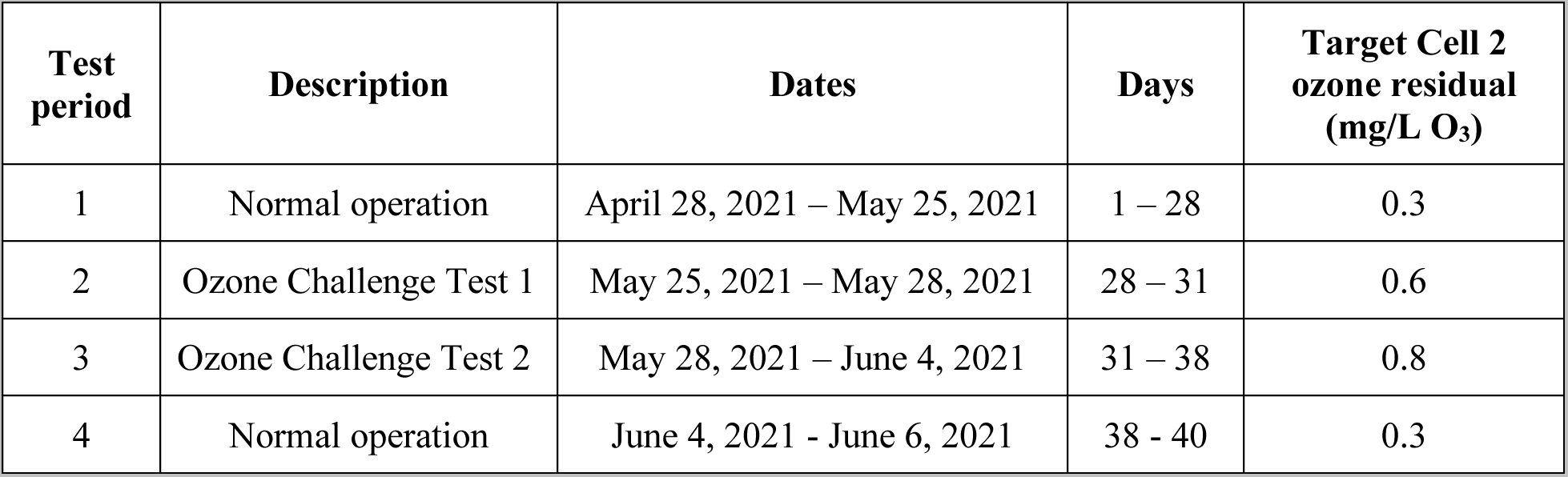
Summary of testing periods and target ozone residuals in ozone contactor Cell 2.

| Test period | Description | Dates | Days | Target Cell 2 ozone residual (mg/L O<sub>3</sub>) |
| --- | --- | --- | --- | --- |
| 1 | Normal operation | April 28, 2021 – May 25, 2021 | 1 – 28 | 0.3 |
| 2 | Ozone Challenge Test 1 | May 25, 2021 – May 28, 2021 | 28 – 31 | 0.6 |
| 3 | Ozone Challenge Test 2 | May 28, 2021 – June 4, 2021 | 31 – 38 | 0.8 |
| 4 | Normal operation | June 4, 2021 - June 6, 2021 | 38 - 40 | 0.3 |

### 2.4 NRT-FCM

The NRT-FCM system consisted of a benchtop flow cytometer (Sysmex Cube 6 V2M, Sysmex USA, Lincolnshire, IL, USA) equipped with one 60 mW, 488 nm laser and five detectors (side scatter, forward scatter, and three fluorescence detectors) and an automated sample preparation system (OnCyt OC-300 Duo, OnCyt Microbiology, Dübendorf, CH; Figure 1). Laser alignment and system calibration were performed at the start of the experiment to verify the system was functioning correctly. Voltages were set at the beginning of the test period using 0.5 µm calibration beads (Sysmex), 3 µm calibration beads (Sysmex), and 3 µm green, fluorescent beads (CountCheck Beads Green, Sysmex). Continuously flowing sample lines supplied water from the ozone contactor influent (Cell 1) and effluent (Cell 7) to the system using sterile, ozone-resistant, flexible tubing (Masterflex, Cole-Parmer, Vernon Hills, IL, USA, cat. no. 644225) and peristaltic pumps (Masterflex, Cole-Parmer). The tubing was inserted into the contactor through six-inch (15.2 cm) diameter stainless steel vent pipes above Cells 1 and 7. Tubing was replaced with new, sterile tubing three times during the study to limit the effect of biofilm growth. Sample lines were run at 75 mL/min, with flow rates checked daily and adjusted as needed.

The automated sampling and staining system collected and processed samples from each location for total and intact cell analyses approximately every 25 minutes, with cleaning cycles between contactor influent and effluent analyses, yielding TCC and ICC results from both locations every 50 minutes. SYTO (1,000x, CyStain^TM^ BacCount Total, Sysmex) was used for TCC, and SYTO and propidium iodide (CyStain^TM^ BacCount Viable, Sysmex) were used for ICC. Each batch of stains was 35 mL and consisted of 70 µL of the stains, 6 to 7 mL of the buffer included in the stain kits (CyStain^TM^ Dilution Buffer, 6.9 mL for TCC and 6.3 mL for ICC), and 28 mL of ultrapure water (UltraPure^TM^ DNase/RNase-Free Distilled Water, Thermo Fisher Scientific, Waltham, MA, USA). Sodium thiosulfate was added to the stain solutions at a concentration of 10 g/L to quench any disinfectant residual in the samples. For both analyses, 350 µL of the stain solution was added to 350 µL of sample. The samples were then incubated with the stains for 10 minutes at 38°C and sent to the flow cytometer, where 100 µL of sample was analyzed. Data loss occurred twice due to user error. The first event occurred on Day 4 and was resolved the morning of Day 5; the second one occurred on Day 26 and was resolved the morning of Day 27.

Two cleaning protocols were used for the NRT-FCM system. During the first eight days of the experiment, the automated sampling and staining instrument and the flow cytometer sample tube and needle were rinsed with 1) a 1% sodium hypochlorite solution, 2) a 50 mM sodium thiosulfate solution, and 3) ultrapure water. The ultrapure water was taken up and analyzed by the cytometer, rinsing the internal components. A more intensive cleaning protocol was initiated on Day 9, consisting of the same protocol, except that each of the three solutions was taken up and analyzed by the cytometer. Additionally, sodium hypochlorite solution and sheath fluid were passed through the automated sampling and staining instrument and cytometer at least weekly to further clean system components.

### 2.5 Real-time physicochemical monitoring

The water treatment plant monitors flow rates, ozone contactor influent pH and temperature, and ozone contactor effluent turbidity in real time. Data were downloaded from the supervisory control and data acquisition (SCADA) system every five minutes. Sensor nodes were used to record real-time oxidation-reduction potential (ORP), pH, electroconductivity, and temperature in the ozone contactor influent and effluent using the continuously flowing sample lines (Figure 1) (Martinez Paz et al., 2022). Every hour, three measurements were recorded for each parameter. Additional details of online monitoring instrumentation are included in Table S1.

### 2.6 Grab sample collection

Grab samples were collected daily from sample tees in the continuously flowing sample lines after the connections to the automated sampling and staining instrument. Samples were collected by flushing the sample line, then collecting one L of water into sterile polypropylene containers. For cATP, impedance FCM, and HPC, 100 mL of sample were transferred to 120 mL sterile containers with excess sodium thiosulfate (IDEXX Laboratories Inc., Westbrook, Maine, USA). The remaining volume was used for pH, temperature, turbidity, and total organic carbon (TOC) analyses. DNA samples were collected two times per week into sterile, 10 L containers using peristaltic pumps and sterile tubing, which was fed into the contactor at the same locations as the continuously flowing sample lines. DNA samples were transported to the laboratory in coolers with cold packs for processing. During the 2022 follow-up testing, HPC samples were collected from the ozone system sample manifold.

### 2.7 Grab sample physicochemical sample analyses

Temperature, pH, turbidity, and TOC were measured daily in triplicate. Temperature and pH were measured using a benchtop probe (Orion^TM^ Versa Star Pro^TM^, Thermo Scientific, USA). Turbidity was measured using a benchtop turbidimeter (Hach TU5200, Loveland, CO, USA). TOC samples were collected in carbon-free 40 mL amber glass vials (Fisher Scientific, Hampton, NH, USA) and acidified to a pH less than 2 using 85% phosphoric acid. Samples were stored at 4°C and analyzed within 28 days. Samples were analyzed using the non-purgeable organic carbon method using a TOC analyzer (Shimadzu TOC-V, Kyoto, JPN). Ozone grab samples were collected from an ozone system sampling manifold and immediately analyzed for ozone residual using a portable spectrophotometer and reagent ampules (Hach Method 8311, Loveland, CO, USA).

### 2.8 Microbial sample analyses

cATP was quantified using the LuminUltra Quench-Gone^TM^ Aqueous Test Kit following manufacturer instructions (LuminUltra Technologies, Ltd., Fredericton, NB, CAN) and a LuminUltra PhotonMaster^TM^ luminometer. Volumes of 50 mL and 100 mL were analyzed for the ozone contactor influent and effluent, respectively. Relative light units (RLUs) were converted to cATP per manufacturer guidance (Equation S6). Analyses were performed in triplicate. HPC analysis was performed using the pour plate method (SM 9215-B-2000)(Standard Methods Committee of the American Public Health Association, American Water Works Association, and Water Environment Federation, 2017). Impedance FCM was conducted using a BactoBox (SBT Instruments A/S, Herlev, DNK) with a 15-minute analysis time and 25 mL sample volumes. Sample conductivity was adjusted using sterile phosphate buffer saline (Gibco, Thermo Fisher Scientific) and ultrapure water as needed to achieve a conductivity within the range of the instrument (500-1,100 microsiemens per cm [µS/cm]). Samples above the upper limit of detection were diluted with ultrapure water. Additional details of analytical methods are included in Table S1.

DNA samples were filtered onto sterile, 0.22 µm polyethersulfone membrane Sterivex^TM^ cartridge filters (MilliporeSigma, Burlington, MA, USA) using sterile tubing and peristaltic pumps. Filtered volumes ranged from 3.2 L to 10.5 L (Table S2). Filters were placed in sterile bags and frozen at −80°C. In addition to the ozone process samples, filter controls (unused cartridge filters) and filtration set-up controls (filtered ultrapure water) were collected during each sampling event. To extract the DNA, filters were aseptically removed from the cartridge casing and transferred to Lysing Matrix E bead tubes (MP Biomedicals, Solon, OH, USA). DNA was extracted from the filters using a modified protocol for the QIAGEN DNeasy PowerWater kit (Hilden, DEU) that includes enzymatic lysis and chloroform-isoamyl (Vosloo et al., 2019). Extraction controls (empty bead tubes) were included in each extraction session. Extracted DNA was quantified using a Qubit 2.0 Fluorometer and the dsDNA High-Sensitivity kit (Thermo Fisher Scientific).

Twenty-five samples were sent for 16S rRNA gene sequencing, including the 22 ozone system samples and three pooled controls (filter controls [n=7], filtration set-up controls [n=7], and DNA extraction controls [n=2]). DNA was amplified using primers targeting the V4 region of the 16S rRNA gene and a dual-indexing sequencing strategy (Kozich et al., 2013). If samples failed to amplify with standard PCR, touchdown PCR was used. Sequencing was performed using an Illumina MiSeq using 250 base pair paired-end reads and the MiSeq Reagent Kit V2 500 cycles (Illumina, San Diego, CA) as previously described (Baker et al., 2021). Library preparation, sequencing, demultiplexing, and primer removal were performed by the University of Michigan Microbiome Core. Samples were named based on the sampling location (INF: ozone contactor influent, EFF: ozone contactor effluent) and the sampling event (from one to 11). Two ozone effluent samples (EFF-4 and EFF-6) were excluded from the analysis due to low read counts (less than 1,000). Total reads across the remaining 23 samples ranged from 5.06×10^4^ to 2.82×10^5^ reads per sample. Raw sequencing data are available from the NCBI Sequence Read Archive (SRA) at BioProject ID PRJNA954821.

### 2.8 Data analysis

The flow cytometer .fcs files (n=4,402) were analyzed to determine total and intact cell concentrations using batch processing in FCS Express 7 (v.7.12.0007, De Novo Software, Pasadena, CA, USA) using previously established fixed gates, which were developed using preliminary data with ozone contactor influent and effluent samples. Dual-gating was employed for all samples to limit the effect of cellular debris and background signal for the intact cell measurements (Figure S2). Outlier analysis was conducted to identify samples impacted by cytometer malfunctions (Figure S3). This analysis was performed using the fluorescence versus time data for each sample with two criteria: 1) samples were flagged if gaps in events were present that were longer than 0.5 seconds, and 2) samples were flagged if, when data from each sample was divided into equally sized bins, the standard deviation of the mean fluorescence for the bins was greater than 0.05. Samples determined to be outliers (n=432) were removed from the dataset. To de-noise the FCM results, the rolling median of three values was calculated and used for all plotting and analyses using the *roll.median* function in the R package *zoo* (Figure S4) (Zeileis and Grothendieck, 2005).

The 16S rRNA gene sequencing reads were demultiplexed and processed using DADA2 following the Pipeline Tutorial using the default settings (version 1.16; https://benjjneb.github.io/dada2/tutorial.html), except that samples were pooled for inference (Callahan et al., 2016). Taxonomy was assigned using the SILVA Release 138.1 training dataset with species assignment using exact matching (McLaren and Callahan, 2021; Quast et al., 2012). The three negative controls and one ozone contactor influent sample (INF-6) that showed evidence of contamination (80% *Actinobacteria* relative abundance and 10% *Ottowia* relative abundance) were excluded from diversity analyses. The 16S rRNA gene sequencing results bar plot and the non-metric multidimensional scaling (NMDS) plot were created using the R package *phyloseq* (v.1.40.0) (McMurdie and Holmes, 2013). Alpha and Beta diversity were calculated using the “diversity” function of the R package *vegan* (v.2.6-2) (Oksanen et al., 2022). Permutational multivariate analysis of variance (PERMANOVA) analysis was performed using the *vegan* “adonis2” function with Bray-Curtis distance. To determine the log2 fold change in relative abundances of specific taxa, the R package *DESeq2* (v.1.36.0) was used (Love et al., 2014).

All other data processing, hypothesis testing, statistical analyses, and plotting were performed using R (version 4.1.1) and RStudio (version 1.4.1717)(R Core Team, 2021; RStudio Team, 2020). R packages used for data analysis include *dplyr*, *readxl*, *readr*, *data.table*, *lubridate*, *scales*, *ggplot2*, *ggpubr*, *stats*, *viridis*, *fuzzyjoin*, *broom*, *devtools*, *zoo*, and *forcats*. Values for results that were less than the lower limit of detection (LLOD) were set to one-half the LLOD for analysis and plotting. Values for results above the upper limit of detection (ULOD) were set at the ULOD (Table S1). To de-noise sensor node data, the rolling median of three values was used for all plotting and analyses (Figure S5). Results of analyses are reported as the median ± one standard deviation unless otherwise stated. Student’s t-tests were used to test the statistical significance of differences between analyses using a threshold for significance of p=0.05. Correlation analyses were performed using linear regression using the “lm” function and Kendall’s tau rank correlation method using the function “cor.test”, both of which are in the R package *stats*. To match daily grab samples and FCM data for combined analyses, FCM measurements that occurred within two hours before or after the collection of a grab sample were averaged and paired with the grab sample result. To match the ozone system data (applied dose, ozone residual, CT), which were collected every four hours, with the FCM data for combined analyses, FCM measurements that occurred within one hour before or after the collection of the sample were averaged and paired with the ozone sample result.

## 3. Results and Discussion

### 3.1 Ozone system performance using surrogates

The ozone system was operated to target Cell 2 ozone residuals ranging from 0.3 to 0.8 mg/L O_3_. During Test Period 1 (Days 1-28), when the target Cell 2 ozone residual was 0.3 mg/L O_3_, the median Cell 2 ozone residual was 0.36 mg/L O_3_ (n=150, Figure S6), the median applied ozone dose was 2.41 mg/L O_3_ (n=150, Figure S7), and the median CT was 0.75 mg-min/L (n=150, Figure S8, Figure 2). To investigate the system’s performance at higher ozone doses, target Cell 2 ozone residuals were then increased starting on Day 28. During Test Period 2 (“Ozone Challenge Test 1”, Days 28-31), the target Cell 2 ozone residual was twice that of Test Period 1 (0.6 mg/L O_3_). The Test Period 2 median Cell 2 ozone residual was 0.77 mg/L O_3_ (n=18), the median applied dose was 2.99 mg/L O_3_ (n=18), and the median CT was 1.52 mg-min/L (n=18). The target Cell 2 ozone residual was then increased to 0.8 mg/L O_3_ for Test Period 3 (“Ozone Challenge Test 2”, Days 31-38) to evaluate system performance at the upper limit of the plant’s ozone dosing. The upper limit of approximately 0.8 mg/L O_3_ was set to limit bromate generation. During this test period, the median ozone residual was 0.91 mg/L O_3_ (n=42), the median applied ozone dose was 2.25 mg/L O_3_ (n=41), and the median CT value was 1.79 mg-min/L (n=42). The system was then returned to normal operation (target Cell 2 ozone residual: 0.3 mg/L O_3_) for Test Period 4 (Days 38-40), and the median Cell 2 ozone residual was 0.41 mg/L O_3_ (n=16), the median applied ozone dose was 2.33 mg/L O_3_ (n=16), and median CT value was 0.57 mg-min/L (n=16).

**Figure 2.**
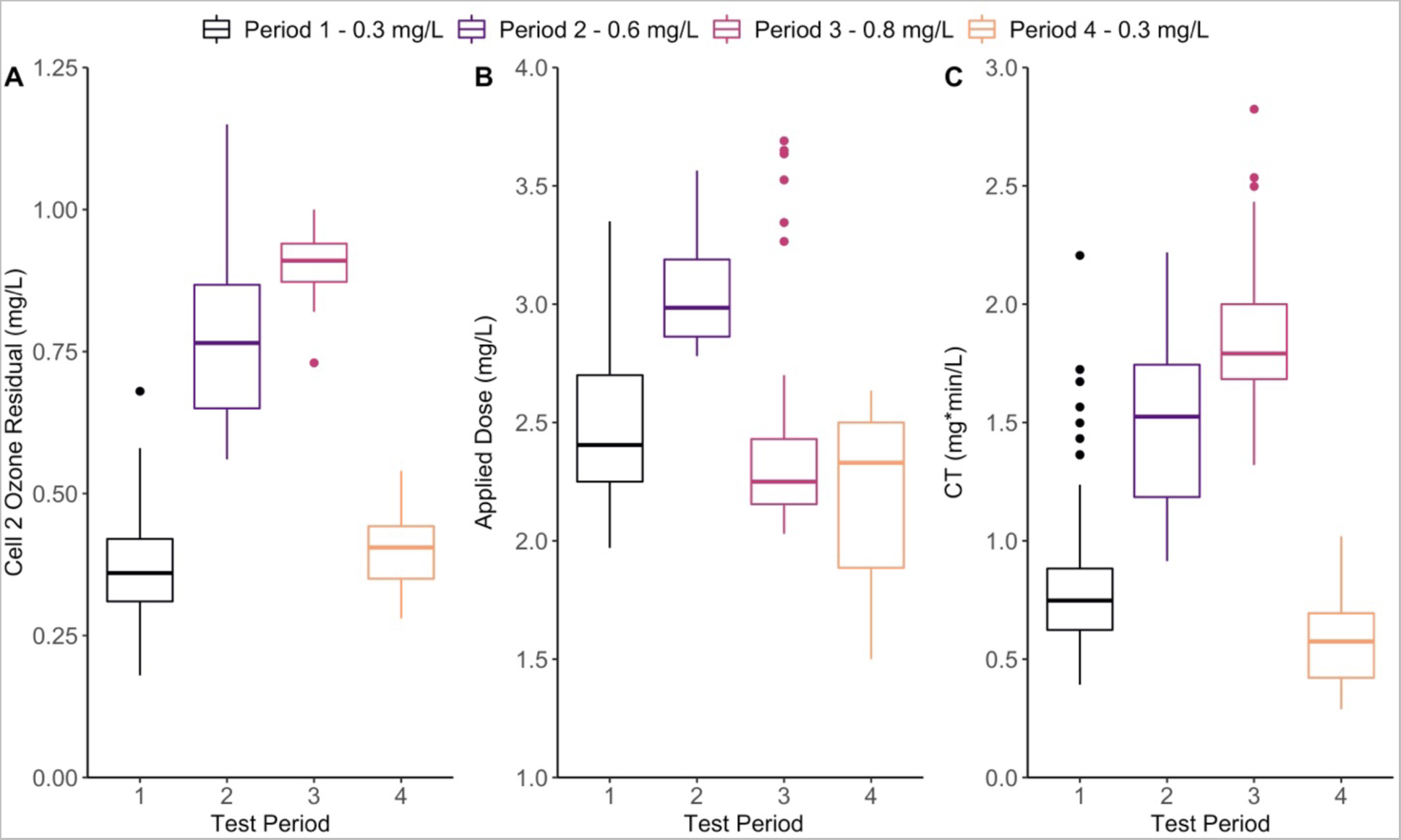
A) Cell 2 ozone residual concentration (mg/L), B) applied ozone dose (mg/L), and C) CT (mg-min/L) over the four test periods. The boxes are colored by test period. Test Periods 1 and 4: 0.3 mg/L O_3_, Test Period 2: 0.6 mg/L O_3_, Test Period 3: 0.8 mg/L O_3_.

The ozone system functioned as expected during the first 24 days of the study. However, on Day 25 of testing, the water treatment plant reported that the ozone contactor was not responding as usual to increases in the ozone gas feed. A lack of response in ozone residual to increased applied ozone doses persisted through the remainder of the study, ultimately leading to the contactor being shut off one week after the study concluded.

### 3.2 Grab and online physicochemical results

In addition to ozone residual monitoring, general physicochemical water quality parameters, including temperature, pH, turbidity, and TOC, were monitored using a combination of grab sampling and sensors. Influent and effluent daily grab sample temperatures ranged from 14.4°C to 21.2°C (median: 17.2°C, n=40) and from 15.2°C to 21.3°C (median: 17.5°C, n=40) and were not significantly different (p>0.05) (Figure S9). Median pH values were higher in the ozone contactor influent (grab sample median: 8.2, n=40) compared to the effluent (grab sample median: 8.1, n=40; p<0.01; Figure S10). Turbidity was measured in grab samples and through an online monitor for the ozone effluent. Influent turbidities ranged from less than 1.0 NTU to 42.7 NTU (median: 7.1 NTU, n= 40; Figure S11). Effluent turbidities were lower, ranging from 0.5 NTU to 17.0 NTU (median: 6.0 NTU, n= 40) in the grab samples, which was significantly lower than in the influent turbidity (p<0.01) and from 1.0 NTU to 28.1 NTU (median: 6.6 NTU, n= 11,447) using the online turbidimeter. Total organic carbon concentrations were similar in the influent (median: 3.5 mg/L, n= 40) and effluent (median: 3.5 mg/L, n= 40; Figure S12).

The periods of high turbidities in the ozone contactor influent confirmed that substantial lime solids carryover occurred during the study. Further, the significantly lower turbidities in the effluent suggested that settling of lime softening solids was occurring in the ozone contactor, which is a known issue in the water treatment plant and was previously described by Kotlarz et al. 2018. For this reason, the contactors are taken out of service and the solids are removed yearly. The contactor used in this study had been offline for one year prior to testing and had not been cleaned prior to the study, suggesting that lime solids were in the contactor when the experiment was initiated. Though few other studies have reported the interference of lime softening solids with ozonation, carryover of lime solids has been reported in filters downstream of softening (United States Environmental Protection Agency, 1999). These complexities of the ozone system made it an ideal location for exploring microbial dynamics with NRT-FCM.

### 3.3 NRT-FCM TCC and ICC quantification

The NRT-FCM system recorded TCC and ICC values in the ozone contactor influent and effluent. Over the course of the experiment, ozone contactor influent TCC and ICC ranged from 1.3×10^5^ to 1.1×10^6^ cells/mL (median: 4.0×10^5^ cells/mL, n=1,044) and from 6.8×10^4^ to 5.8×10^5^ cells/mL (median: 2.3×10^5^ cells/mL, n=958), respectively (Figure 3). While the influent TCC and ICC were relatively stable during the study, greater variability was observed in the ozone contactor effluent with TCC ranging from 1.9×10^3^ to 3.0×10^6^ cells/mL (median: 1.3×10^5^ cells/mL, n=988), and ICC ranging from 5.6×10^2^ to 7.9×10^5^ cells/mL (median: 4.2×10^4^ cells/mL, n=972). During the first three days of the experiment, effluent TCC and ICC were lowest, with median values of 1.5×10^4^ cells/mL (n=70) and 1.5×10^3^ cells/mL (n=74), respectively. The median effluent TCC and ICC increased from Days 4 through 20 to 1.7×10^5^ cells/mL (n=401) and 4.2×10^4^ cells/mL (n=395), respectively. Compared to the Test Period 1 median influent TCC (4.4×10^5^ cells/mL, n=679) and ICC (2.4×10^5^ cells/mL, n=616), the median influent TCC and ICC during Test Period 2 (median TCC: 3.0×10^5^ cells/mL, n=85; median ICC: 1.3×10^5^ cells/mL, n=75) and Test Period 3 (median TCC: 3.6×10^5^ cells/mL, n=200; median ICC: 2.2×10^5^ cells/mL, n=187) were slightly lower. The effluent TCC was also lower during Test Period 2 (median TCC: 7.4×10^4^ cells/mL, n=84) compared to Test Period 1 (median TCC: 1.1×10^5^ cells/mL, n=646), but increased to above Test Period 1 levels in Test Period 3 (median TCC: 1.5×10^5^ cells/mL, n=187). During the final test period (Test Period 4), the effluent TCC (4.5×10^5^ cells/mL, n=71) and ICC (3.3×10^5^ cells/mL, n=71) exceeded the influent TCC (3.8×10^5^ cells/mL, n=80) and ICC (2.5×10^5^ cells/mL, n=80). Effluent TCC and ICC were similar to or exceeded the corresponding influent cell counts on a few occasions throughout the study.

**Figure 3.**
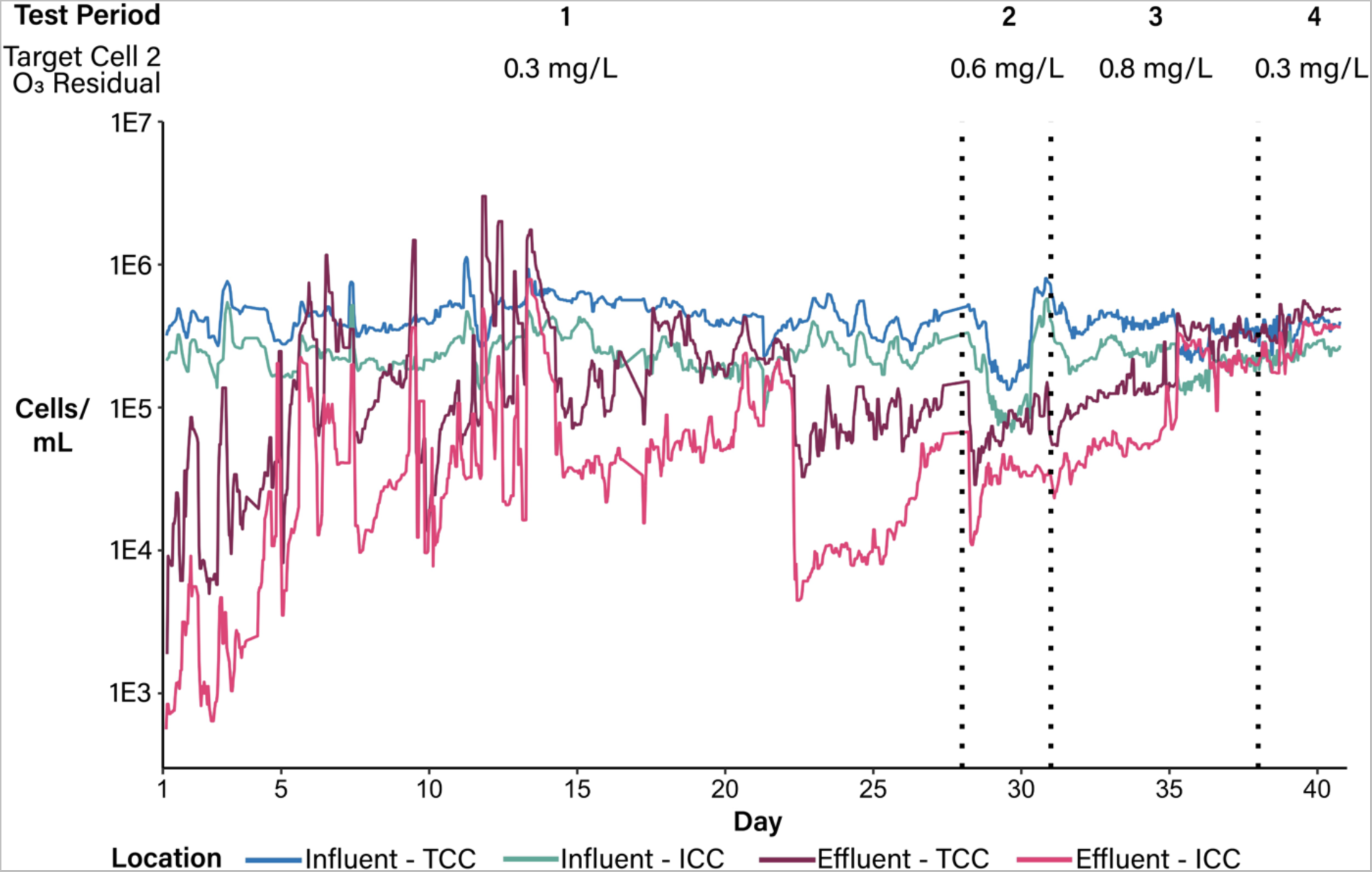
Total cell counts (TCC) and intact cell counts (ICC) in the ozone contactor influent and effluent, as determined using near real-time flow cytometry. Data are plotted as the rolling median of three measurements. Vertical black dotted lines indicate the test periods, during which different Cell 2 ozone residuals were targeted: Test Periods 1 and 4: 0.3 mg/L O_3_, Test Period 2: 0.6 mg/L O_3_, Test Period 3: 0.8 mg/L O_3_.

Net log removal (contactor effluent vs. contactor influent) of intact and total cells over the course of the experiment also varied, reflecting the changing effluent cell concentrations (Figure 4). Overall, the median log removal of intact cells was 0.68 (n=776), and the median log removal of total cells was 0.48 (n=776). Log removal values were highest during the first three days of the experiment (median log removal TCC: 1.39, n=17; median log removal ICC: 2.25, n=16), with the maximum log removal of total cells occurring on Day 1 (2.25) and the maximum log removal of intact cells occurring on Day 3 (2.56; Figure S13). However, log removal values began to decrease after Day 3 despite consistent ozone dosing, including periods where total and intact cell log removals were near zero or negative. Despite substantially higher CT values during Test Periods 2 and 3, log removal values similar to those observed during the first days of the study were not achieved, with maximum log removals after Test Period 1 occurring on Day 28 (Test Period 2) for both total (1.14) and intact (1.24) cells. By Test Period 4, the maximum net log removal of intact cells was 0.1, and the median log removal was negative (−0.1, n=16).

**Figure 4.**
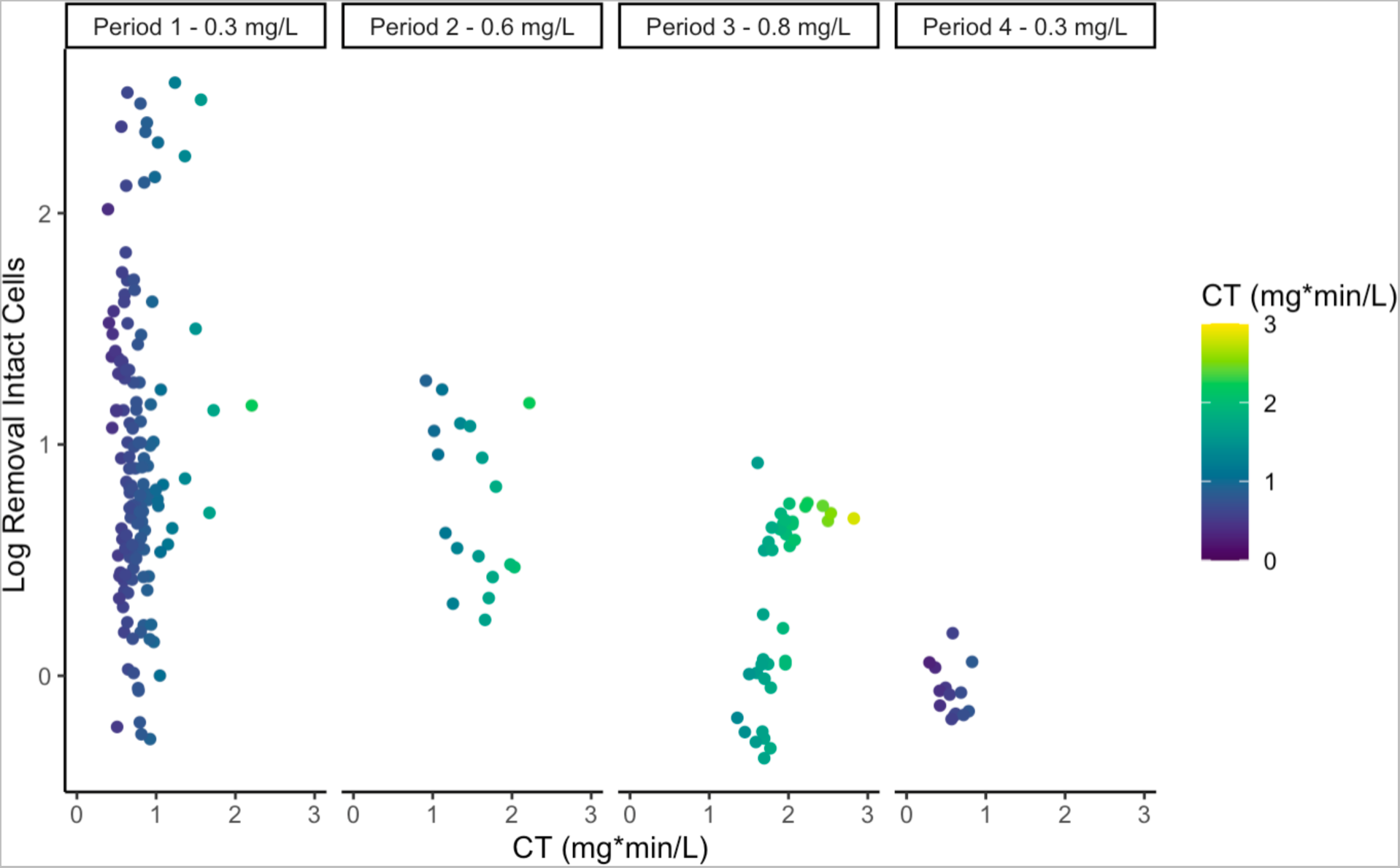
Net log removal of intact cells using NRT-FCM versus CT, grouped by test period. Points are colored by CT. Influent and effluent cell concentrations used to calculate log removal are the mean of all measurements taken within one hour before or after ozone residual grab sampling.

Though monitoring of ozone residual, CT, and applied dose facilitate the maintenance of a minimum level of disinfection and thus compliance with regulatory requirements, they may lack sensitivity in terms of determining contactor effluent water quality. In this study, the ozone system was relatively stable and performing as expected until Day 25, when an issue with ozone residuals was noted. However, the NRT-FCM revealed substantial fluctuations in contactor effluent over time, with increased effluent cell concentrations found just days after the system was placed in service. Correlation analysis using Kendall’s tau-b to evaluate the relationship between CT and log intact cell removal showed that, while the correlation is statistically significant (p=0.04), the correlation coefficient is small (Kendall’s tau=-0.091). The relationship between log intact cell removal and Cell 2 ozone residual was also statistically significant (p=0.01, Kendall’s tau=-0.11). Log total cell removal did not significantly correlate with CT (p=0.30, Kendall’s tau=0.05) or Cell 2 ozone residual (p=0.50, Kendall’s tau=-0.03).

The CT required for inactivation of microorganisms with ozone varies widely. Based on the CTs, which ranged from 0.29 to 2.8 mg-min/L, and water temperatures, which ranged from 14 to 21 °C, log removal credits for *Giardia* spp. ranged from 0.5 log to over 3 log, <0.25 log to 0.5 log for *Cryptosporidium* spp, and up to 4 log for viruses (US EPA, 2010b, 1999). For bacteria, these CTs would achieve substantial inactivation of susceptible species, such as *E. coli*, but little inactivation of resistant species, such as the spores of *Bacillus subtilis* (Hunt and Mariñas, 1997; Larson and Mariñas, 2003). As no quantification of regulated pathogens such as *Cryptosporidium* spp., viruses, or *Giardia* spp. was conducted during this study, inactivation of specific microorganisms was not assessed. Additionally, the system is only permitted for 0.5 log inactivation of *Giardia* spp. and 2 log virus removal, not for *Cryptosporidium* spp. However, previous studies have reported that the CT guidance developed by the US EPA is conservative and effective for achieving removal of target microorganisms through ozonation (Morrison et al., 2022).

Previous studies have identified several potential causes of outliers in FCM datasets, including cytometer clogging, changes in pressure or sample flow rate, bubbles, and large particles, and a number of methods have been developed to identify outliers in FCM datasets (Fletez-Brant et al., 2016; Le Meur et al., 2007; Meskas et al., 2022; Monaco et al., 2016). In this study, issues relating to potential microbial growth in the system, inconsistent sample flow rates, and occasional deviations from the target sample volume were encountered. On two occasions (Days 14 and 23), effluent cell counts dropped substantially (>1 log) and were not linked to similar decreases in influent cell counts. This is likely linked to intensive cleaning events (Figure S14), where the tubing was replaced with sterile tubing, and the NRT-FCM system was thoroughly cleaned by flushing the system with bleach, sodium thiosulfate, and sterile water. Microbial growth in the flexible tubing used to feed the NRT-FCM system may also have contributed to this variation. Further research is needed to investigate materials and system designs that will limit the potential for microbial growth through NRT-FCM systems. Interruptions or changes in the flow of the sample through the cytometer (Figure S3A) and jumps or shifts in the fluorescence of the events during sample analysis (Figure S3B) also occurred, likely due to common malfunctions of the flow cytometer, which were flagged as outliers and removed from the data set (Meskas et al., 2022). Additionally, the cytometer occasionally analyzed more than 100 µL of sample. Each time this occurred, the system was restarted and returned to normal operation. These events did not significantly impact measurements (Figure S15). After the study, an issue with the automated sampling and staining system code was identified that caused non-ideal mixing of the SYTO stain in TCC samples. We determined that this issue may have reduced TCC by approximately 10%. However, as this would not significantly impact findings, the decision was made not to adjust results. In future studies, continuous quality control checking and analysis of NRT-FCM data will be needed. Automation of quality control and data analysis will also facilitate the broader adoption of NRT-FCM in the drinking water industry.

### 3.4 Relationship between NRT-FCM and grab microbial parameters

Daily grab samples were analyzed for cATP, HPC, and intact cells using impedance FCM. cATP concentrations ranged from 2.3 to 75 pg cATP/mL in the influent (median: 9.2 pg cATP/mL, n=39) and from <0.1 to 27 pg cATP/mL in the effluent (median: 3.6 pg cATP/mL, n=39; Figure S16A). HPC ranged from 4.0×10^2^ to 5.1×10^4^ CFU/mL (median: 4.4×10^3^ CFU/mL, n=40) in the influent and 2.0 ×10^3^ to 6.6×10^3^ CFU/mL (median: 2.1×10^3^ CFU/mL, n=40) in the effluent (Figure S16B). Results for impedance FCM ranged from 9.0×10^2^ to 3.0×10^4^ intact cells/mL (median: 3.5×10^3^ intact cells/mL, n=40) in the influent and 5.7×10^2^ to 1.4×10^4^ intact cells/mL (median: 2.1×10^3^ intact cells/mL, n=40; Figure S16C) in the effluent.

The relationships between the daily microbial grab samples and the NRT-FCM results were investigated using linear regression and Kendall’s tau. The linear regression analysis for cATP, HPC, and impedance FCM and intact cells from NRT-FCM showed positive correlations of varying strengths (Figure S17). The microbial measure that showed the highest correlation with ICC was cATP, with a linear regression R-squared value of 0.47. The HPC and impedance FCM results also positively correlated with ICC, but the correlations were weaker (HPC R-squared: 0.33; impedance FCM R-squared: 0.19). Similar results were found when evaluating correlations using Kendall’s tau-b, with the strongest correlation being between ICC and cATP (Kendall’s tau: 0.40, p<0.05), followed by impedance FCM (Kendall’s tau: 0.30, p<0.05) and HPC (Kendall’s tau: 0.20, p<0.05).

Net log removals for the daily microbial grab sample also varied over the course of the experiment, and similar trends in log removal were observed in the cATP, HPC, and impedance FCM data (Figure S18). To investigate the differences in log removal values between the daily grab samples and NRT-FCM, median log removal values were compared (Figure 5). Compared to the daily microbial grab samples, the net log removal of intact cells using NRT-FCM was significantly higher (p<0.05) than that observed with HPC, cATP, and impedance FCM. The highest median log removal for the daily grab sample microbial analyses was 0.56 (n=39) using cATP. The median log removal for HPC and impedance FCM were lower at 0.31 (n=40). The median daily matched log removal for intact cells using NRT-FCM was 0.75 (n=40). The median daily log removal for NRT-FCM total cells was significantly lower than for intact cells at 0.38 (n=40; p<0.05). For total cell log removal, the log removal values were not significantly different from the HPC or the impedance FCM log removals (p>0.05) but were significantly lower than the cATP log removal (p<0.05).

**Figure 5.**
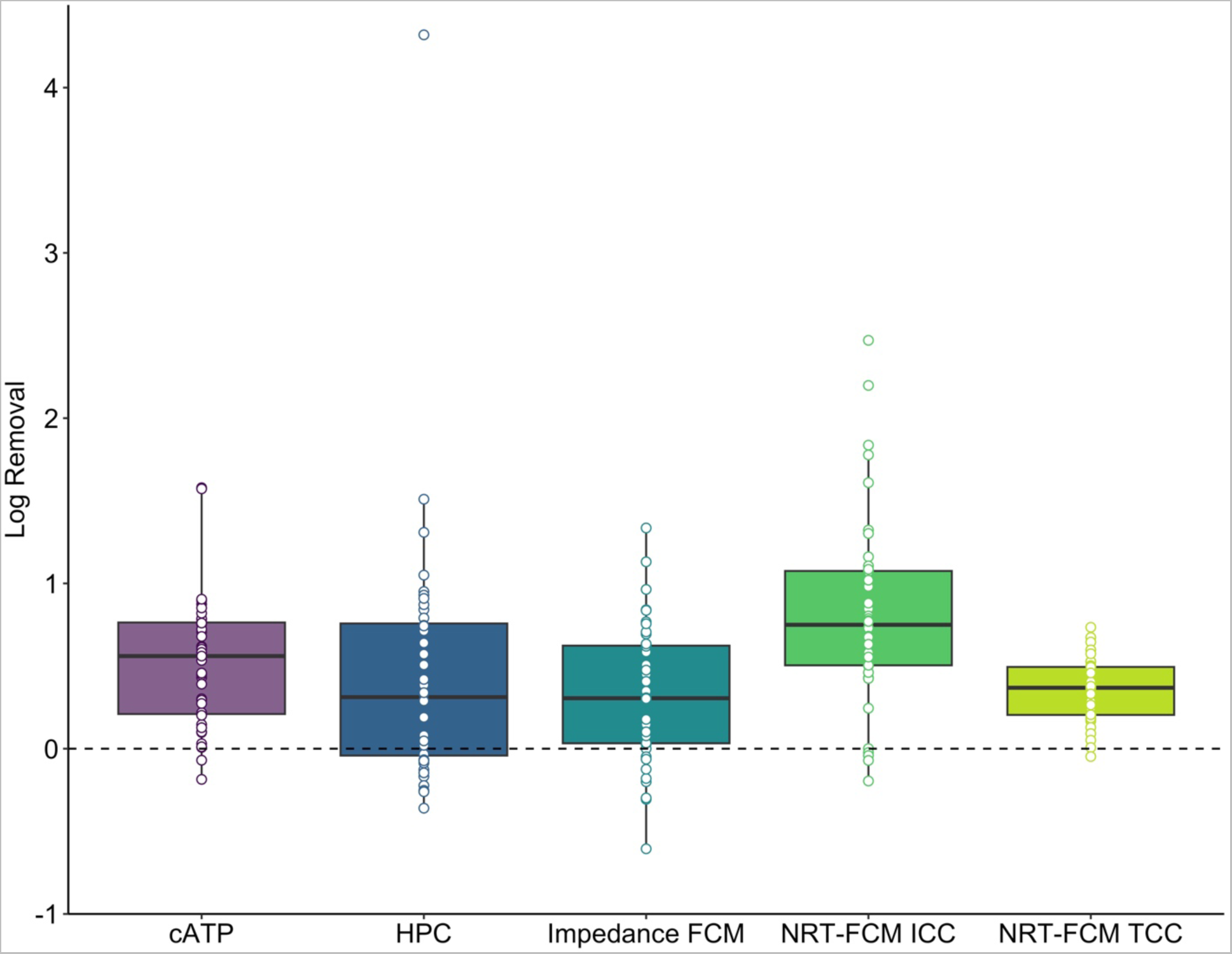
Boxplots showing the net log removal values for the daily microbial analyses (cellular adenosine triphosphate [cATP], heterotrophic plate counts [HPC], and impedance flow cytometry [impedance FCM]), and the near real-time flow cytometry (NRT-FCM) intact cell counts (ICC) and total cell counts (TCC) for samples collected from the contactor influent and effluent. Daily NRT-FCM log removals are the mean of all measurements two hours before and after grab sample collection.

The correlations between ICC and the daily grab sample results were positive but relatively weak, ranging from Kendall’s tau values of 0.2 for HPC to 0.47 for cATP. The relationship between HPC and TCC was similar to previous reports in the literature, with HPC being approximately two log lower than TCC values, demonstrating the large difference in results when cells are quantified with HPC versus FCM (Gabrielli et al., 2021; Van Nevel et al., 2017). The weak correlation between FCM results and HPC agrees with previous studies, which have reported either weak or no correlation between the methods (Cheswick et al., 2022; Van Nevel et al., 2017). cATP showed the strongest correlation to ICC, though the correlation coefficient was less than previously reported (Nescerecka et al., 2014). A potential reason for this may be differences in the methods, as a different cATP method was used, and SYBR green was used instead of SYTO.

### 3.5 Investigation of the microbial community using 16S rRNA gene sequencing

The 16S rRNA gene sequencing results showed that microbial communities in the ozone contactor influent and effluent were substantially different. The total number of amplicon sequence variants (ASVs) in the contactor influent ranged from 1,889 to 3,381 (median: 2,661, n=10). In contrast, total ASVs in the effluent samples ranged from 109 to 2,534 (median: 643, n=9). Rarefaction curves were generated to investigate the observed ASVs by sampling depth for each sample (Figure S19). Dominant phyla in the influent samples included Bacteroidota, Proteobacteria, and Actinobacteriota, while in the effluent samples dominant phyla included Proteobacteria, Cyanobacteria, and Planctomycetota (Figure 6; Table S3). While most ASVs decreased in relative abundance after ozonation, the relative abundance of some ASVs increased, including ASVs assigned to Sphingobacteriales (log2 fold change: 23), *Hyphomicrobium* (log2 fold change: 23), *Gemmata* (log2 fold change: 22), and *Reyranella* (log2 fold change: 21). A previous study that characterized sludge and biofilm samples from the same ozone system using 16S rRNA gene sequencing found that operational taxonomic units (OTUs) identified as *Acidobacteria* subgroup Gp4, *Limnobacter*, Bradyrhizobiaceae, *Brevundimonas*, Rhodobacteraceae, *Herminiimonas*, and *Methylophilus* were among the taxa with highest relative abundances (Kotlarz et al., 2018). These taxa were also detected in the water samples collected in this study, except for *Acidobacteria* Gp4. However, their relative abundances were low (median relative abundances below 0.01%) apart from *Limnobacter*, which had influent and effluent median relative abundances of 0.02% and 0.51%, respectively.

**Figure 6.**
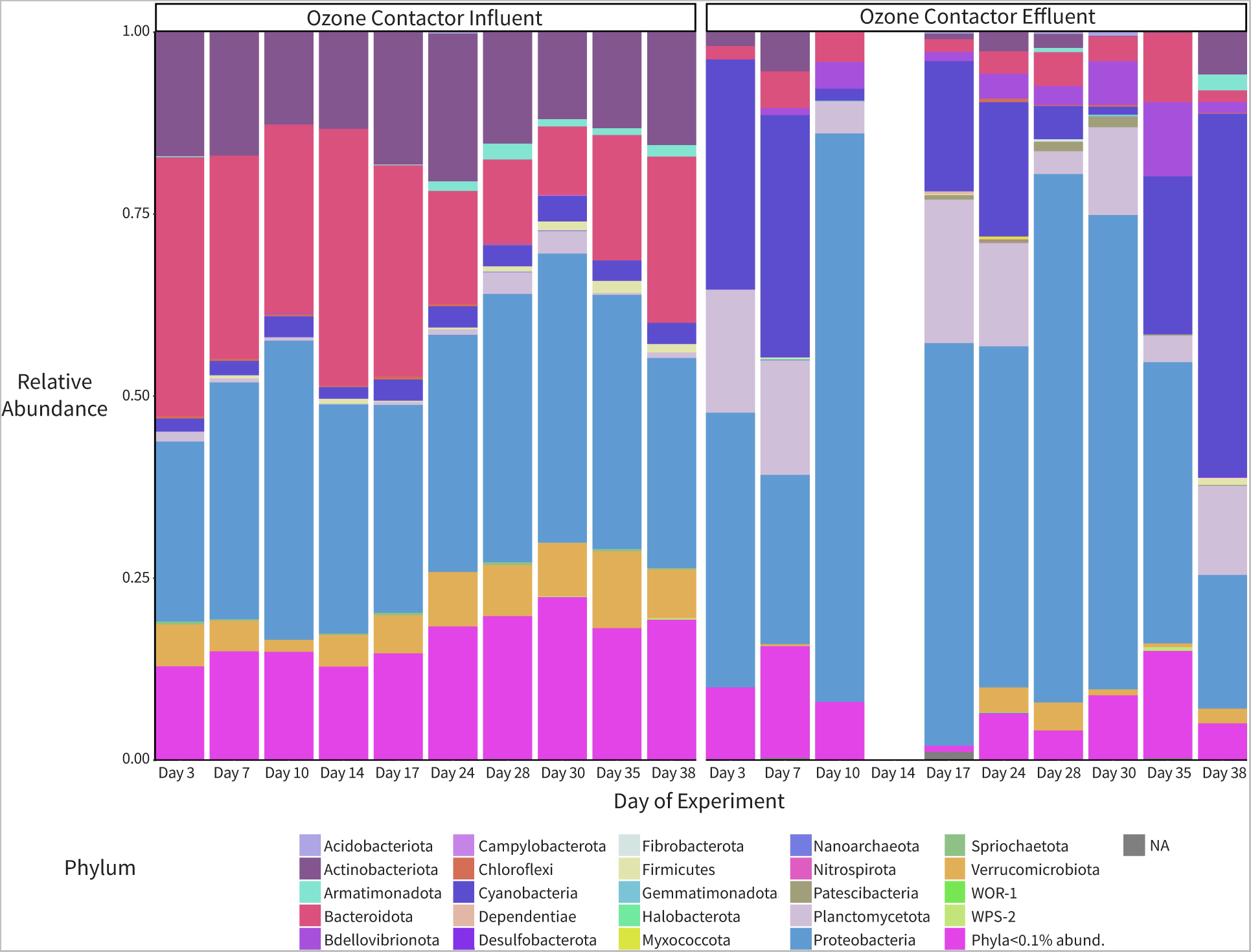
16S rRNA gene sequencing results for the ozone contactor influent (left) and effluent (right) samples at the phyla level. All phyla with at least 0.1% relative abundance are displayed. Samples were collected during 11 sampling events. NA: not classified to the phylum level. The effluent samples for Day 14 (EFF-4) and 21 (EFF-6) were excluded from the analysis due to low read counts (less than 1,000 reads). The influent sample from Day 21 (INF-6) was excluded due to evidence of contamination (80% *Actinobacteria* relative abundance and 10% *Ottowia* relative abundance).

An analysis of four alpha diversity measures (observed ASVs, Chao1, Shannon, and Simpson) showed that, regardless of method, alpha diversity was significantly higher (p<0.05) in the influent compared to the effluent samples (Figure S20). A PERMANOVA analysis indicated that the microbial community compositions in the influent and effluent samples were dissimilar (p=0.001) as visualized by a non-metric multidimensional scaling (NMDS) plot (Figure S21). The influent samples clustered closely together, whereas the effluent samples were less clustered in the NMDS plot, and no clustering pattern by sampling date was apparent. No or weak correlations were observed between the alpha diversity of the effluent samples and log intact and log total cell removals (log intact cell removal: Shannon R^2^ = 0.01, Simpson: R^2^ = 0.03; log total cell removal: Shannon R^2^ =0.05, Simpson: R^2^ = 0.24), indicating that alpha diversity did not significantly change with changes in total or intact cell removal.

The 16S rRNA gene sequencing data were used to investigate the change in the relative abundances of genera that contain bacterial pathogens through the ozone contactor. No *Escherichia* ASVs were found, and two ASVs of the genus *Enterococcus* were found. However, these ASVs were only detected in specific samples (five influent samples and two effluent samples), and relative abundances were low (influent and effluent medians: <0.01%). ASVs of the genera *Legionella*, *Pseudomonas*, and *Mycobacterium*, which contain opportunistic pathogen species, were more prevalent. The total relative abundances of ASVs identified as *Legionella* ranged from 0.14% to 1.5% in the contactor influent (median: 0.3%) and <0.01% to 0.25% in the effluent (median: 0.08%). The total relative abundances of ASVs identified as *Legionella* were significantly higher in the contactor influent samples compared to the contactor effluent samples (p<0.05). For *Pseudomonas*, total relative abundances in the contactor influent ranged from 0.25% to 1.2% (median: 0.62%) and from 0.06% to 24.5% in the effluent (median: 0.56%). Although one effluent sample contained a high relative abundance (24.5%) of *Pseudomonas*, overall, the differences in relative abundances between the influent and effluent were not statistically significant (p>0.05). The total relative abundances of ASVs corresponding to *Mycobacterium* ranged from 0.02% to 0.1% in the contactor influent samples (median: 0.03%) and from <0.01% to 1.0% in the contactor effluent samples (median: 0.16%). Differences in total relative abundances for *Mycobacterium* ASVs between the contactor influent and contactor effluent samples did not meet the threshold for statistical significance (p=0.06).

The 16S rRNA gene sequencing results showed that ozonation caused substantial changes to the contactor effluent microbial community despite periods of low net log removal of cells. We hypothesize that the low TCC and ICC net log removal values observed were the result of the regrowth or release of microorganisms in the later chambers of the contactor, where ozone is not applied and ozone residuals are low or non-detect.

This hypothesis is supported by the results of Kotlarz et al. 2018, who reported that biofilms were present on the contactor walls and on the lime solids sludge that had accumulated at the bottom of the contactor. These biofilms were found to contribute to increased ICC in the effluent (Kotlarz et al., 2018). Unlike the previous study, this study found that effluent ICC increased after contactor start-up, sometimes exceeding influent ICC, suggesting that the duration of ozone contactor operation influences results obtained with discrete microbial sampling. The prevalence of this problem is unclear in the current literature. However, it is suspected that other systems with high concentrations of solids in the ozone contactor influent may also experience sludge accumulation and biofilm growth. For example, Vital et al., 2012 investigated changes in TCC and ICC through water treatment systems in the Netherlands. They reported a log ICC reduction of approximately one through ozonation at one of the facilities. At this facility, the cATP reduction after ozonation was less than one log. The authors hypothesized that higher than expected effluent ICC and cATP values might be due to regrowth after the ozone residual has dissipated. Taken together, the concern of regrowth in the later stages of contactors warrants further investigation.

### 3.6 Follow-up investigations

The NRT-FCM experiment was initiated two days after the ozone train was turned on, after previously being off for approximately five months. The contactor underwent an initial start-up phase during the first few days of operation, where effluent cell concentrations were generally low, after which effluent cell concentrations were higher. Shortly after the experiment concluded, the utility shut down the contactor due to a suspected leak in the ozone gas system. This could have caused the higher than expected ozone residuals in Cell 2 and lower than expected residuals in Cell 3. The utility repaired and placed the contactor back into service in July 2021. Influent and effluent samples were collected and analyzed for HPC in summer 2022 (Figure S22). The median influent HPC was 810 CFU/mL (n=13), and the median effluent result was 2,100 CFU/mL (n=13). The median HPC log removal was −0.25, confirming that regrowth was likely occurring in the contactor despite the cleaning and repairs. This investigation highlights the value of direct microbial quantification of disinfection system performance, particularly for identifying non-ideal performance.

### 3.7 Implications and future work

Automated measurement of microbial parameters offers utilities the ability to quickly make decisions and gauge the performance of drinking water treatment systems. Given the importance of effective disinfection for the reduction of pathogens, automated monitoring is particularly attractive for disinfection systems. Previous studies have reported that NRT-FCM can effectively detect process changes (Besmer and Hammes, 2016) or contamination events (Favere et al., 2021). In this study, an increase in contactor effluent cell concentrations was detected by NRT-FCM but was not detected by surrogates. Ozonation provided an ideal test case for the NRT-FCM system due to the way ozone interacts with bacterial cells, rapidly causing damage to nucleic acids and cell membranes (Hunt and Mariñas, 1999, 1997; Lee et al., 2016), which can be readily observed with FCM using membrane integrity stains (Al-Hashimi et al., 2015; Ramseier et al., 2011). FCM and NRT-FCM also have the potential to be used to monitor chlorine disinfection, as reported earlier (Cheswick et al., 2022, 2020). However, the NRT-FCM system may be less ideal for disinfection systems that result in delayed damage to cell membranes, such as UV. Alternative stains that assess viability based on the activity of cellular processes may facilitate the use of NRT-FCM for UV in the future (Chiang et al., 2021; Grossi et al., 2018; Safford and Bischel, 2019).

While NRT-FCM has shown substantial promise for continuous monitoring of microorganisms in drinking water, water reuse, and wastewater, several challenges remain for its broader implementation in the water industry. Among these are the high troubleshooting and maintenance requirements. The ozone system selected for this study allowed for the evaluation of NRT-FCM in a system with high turbidity. While the system yielded valuable insights into the operation of the ozone system, it required significant maintenance, including daily evaluations and cleaning, priming, or troubleshooting, which occurred at least two times per week. Such a high level of maintenance for a single instrument may not be feasible for water treatment plant staff. However, as NRT-FCM technologies are refined and implemented in more full-scale studies, maintenance requirements will likely be reduced. Certain flow cytometer models currently available offer decreased flexibility in terms of sample analysis but lower maintenance compared to the instrumentation used in this study, which may be preferred for utilities with staffing limitations (Sadler et al., 2020; Gabrielli et al., 2022). Additionally, machine learning technologies may facilitate the automation of data processing and outlier and event detection (Sadler et al., 2020; Kyritsakas et al., 2023). Other factors that may limit the growth of this technology include the high capital cost relative to other microbial monitoring methods and the lack of broad acceptance of FCM, though some utilities and regulators are exploring the use of FCM for monitoring (Safford and Bischel, 2019).

The results of this work highlight how in-depth microbial characterization of water treatment processes can provide insights beyond what can be discerned with physicochemical parameters. The observed increased contactor effluent cell concentrations indicated a potential concern that could be investigated by the water treatment plant. Further, complementing NRT-FCM with 16S rRNA gene sequencing provided additional information for understanding the ozone contactor dynamics. These methods may be particularly valuable for other water treatment plants with suspected non-ideal disinfection systems. Additional work is needed to assess how best to evaluate performance and optimize ozone systems if regrowth is suspected. As microbial monitoring technologies become more affordable and user-friendly, they will facilitate a deeper understanding of full-scale drinking water system operation, with tangible benefits for both drinking water utilities and consumers.

## 4. Conclusions

● NRT-FCM revealed increased contactor effluent total and intact cells over time.
● Of the grab sample microbial analyses, cATP correlated best with NRT-FCM results. The highest net log removal was found using NRT-FCM intact cells; the log removals determined using HPC and impedance FCM were similar to those observed using NRT-FCM total cells.
● 16S rRNA gene sequencing supported the hypothesis that low overall log removals were linked to bacterial regrowth rather than inefficacy of disinfection.
● Equipment and management challenges were encountered with the implementation of the NRT-FCM system at full-scale.
● NRT-FCM may be particularly valuable for the early detection of deviations from typical disinfection system performance.

## Data Availability

Raw sequencing data are available from the NCBI Sequence Read Archive (SRA) at BioProject ID PRJNA954821.

## Acknowledgments

The authors would like to thank the staff of the City of Ann Arbor Water Treatment Plant for their support of this work, Daniela Tapia Pitzzu for help with sampling, and Frederik Hammes for assistance with study design and manuscript review. Funding was provided by the Blue Sky Initiative (College of Engineering, University of Michigan). KSD was supported by a National Science Foundation Graduate Research Fellowship [grant number DGE-1256260] and a University of Michigan Rackham Predoctoral Fellowship. This research was supported by work performed by The University of Michigan Microbiome Core.

## Supplemental Information

Supplementary materials are available in the Supplementary Information file.

