## Supplementary Information for "Assessing the effectiveness of near real-time flow cytometry in monitoring ozone disinfection in a full-scale drinking water treatment plant"

**Overview**

1. Supplementary Figures
2. Supplementary Tables
3. Supplementary Equations
4. Supplementary Text
5. Supplementary References

**Contents**

**1. Supplementary Figures**


- Figure S1. Plant flow during the experiment in gallons per day (gallons/day) and cubic meters per day ($m^3$/day).
- Figure S2. Typical flow cytometry event plots, with red fluorescence (CyStain Red) plotted versus green fluorescence (CyStain Green) using the dual gating strategy.
- Figure S3. Flow cytometry events versus time plots (red fluorescence, "CyStain Red", versus time in seconds) showing examples of outlier data points resulting from changes in sample flow rate in the flow cytometer during analysis.
- Figure S4. Flow cytometry results obtained as described in Figure S2 (blue) and using a rolling median smoothing approach (red), where the median was calculated for three measurements.
- Figure S5. Sensor box pH results (blue) and the same data presented using a rolling median smoothing approach (three measurements; red) for the ozone contactor influent and effluent.
- Figure S6. Ozone contactor Cell 2 ozone residual over time during the four test periods.
- Figure S7. Applied ozone dose over time during the four test periods.
- Figure S8. Concentration x time (CT) over time during the four test periods.
- Figure S9. Temperature results for the ozone contactor influent and effluent using grab sampling, the water treatment plant's sensor, and the sensor boxes.
- Figure S10. pH results for the ozone contactor influent and effluent using grab sampling, the water treatment plant's sensor, and the sensor boxes.
- Figure S11. Turbidity results for the ozone contactor influent and effluent using grab sampling and the water treatment plant's sensor.
- Figure S12. Total organic carbon results for the influent and effluent over the study period.
- Figure S13. Total and intact cell net log removal over time during the four test periods.
- Figure S14. Impact of intensive cleaning events on the NRT-FCM results in the A) ozone contactor influent and B) effluent.
- Figure S15. Impact of volume deviations during testing on the NRT-FCM results.

- Figure S16. Daily microbial grab sample results over time.
- Figure S17. Scatter plots of near real-time flow cytometry intact cell counts (ICC) per mL versus A) cellular adenosine triphosphate (cATP), B) heterotrophic plate counts (HPC), and C) impedance flow cytometry (impedance FCM).
- Figure S18. Net log removal over time for the daily microbial grab samples.
- Figure S19. Rarefaction curves for 16S rRNA gene sequencing results showing the number of amplicon sequence variants (ASVs) versus the number of reads for each sample.
- Figure S20. Alpha diversity results (observed, Chao1, Shannon, and Simpson) for the ozone contactor influent and effluent across the 11 sampling events.
- Figure S21. Results of non-metric multidimensional scaling (NMDS) analysis for the 16S rRNA gene sequencing results.
- Figure S22. Heterotrophic plate count (HPC) results during 2022 follow-up testing.

**2. Supplementary Tables**

- Table S1. Sample analysis methods
- Table S2. Filtered volumes for DNA analyses
- Table S3. 16S rRNA sequencing top 30 ASVs and relative abundances as a fraction of one

**3. Supplementary Equations**

- Equations S1. T10 calculation
- Equations S2. Cell 2 average ozone residual and CT calculations
- Equations S3. Cell 3 average ozone residual and CT calculations
- Equations S4. Cell 4 CT calculation
- Equations S5. Applied ozone dose calculation
- Equation S6. Conversion from relative light units (RLU) to cellular ATP (cATP) per mL of sample

**4. Supplementary Text**

- Supplementary Text 1. Concentration x Time calculations summary
- Supplementary Text 2. Flow cytometry gating strategy summary
- Supplementary Text 3. Cellular ATP sample analysis summary

#### 1. Supplementary Figures

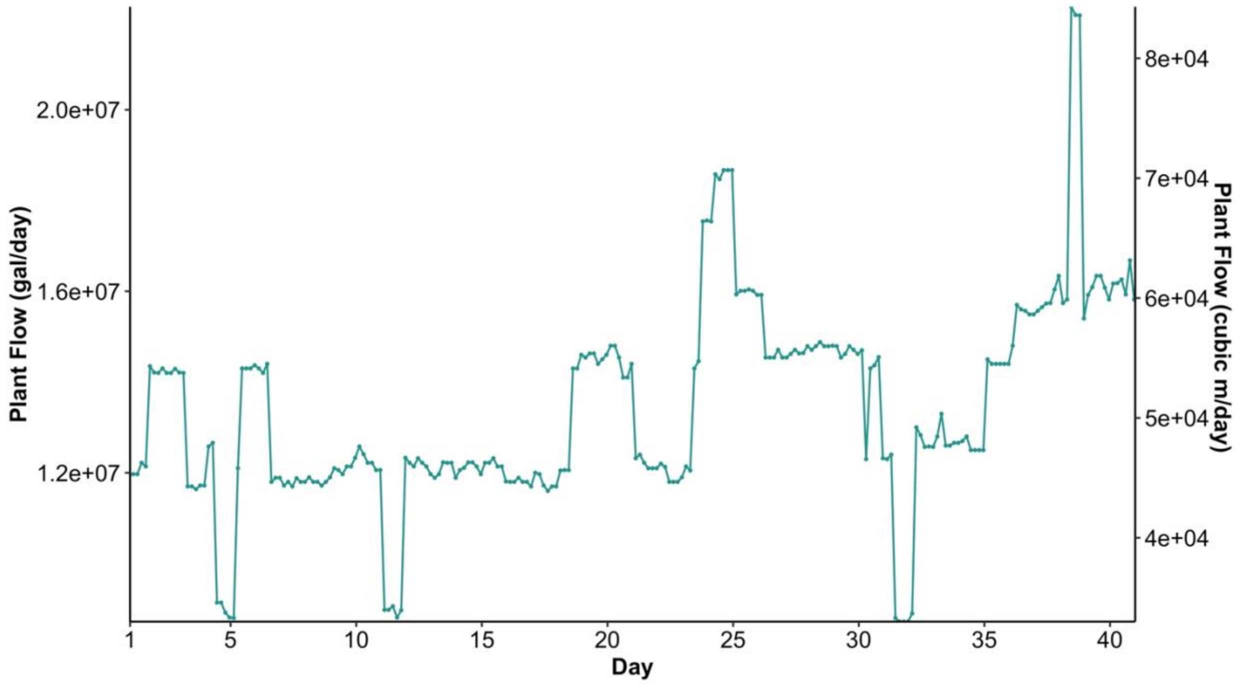


**Figure S1.** Plant flow during the experiment in gallons per day (gallons/day) and cubic meters per day ($m^3$/day). The study took place from April 28, 2021 to June 6, 2021.

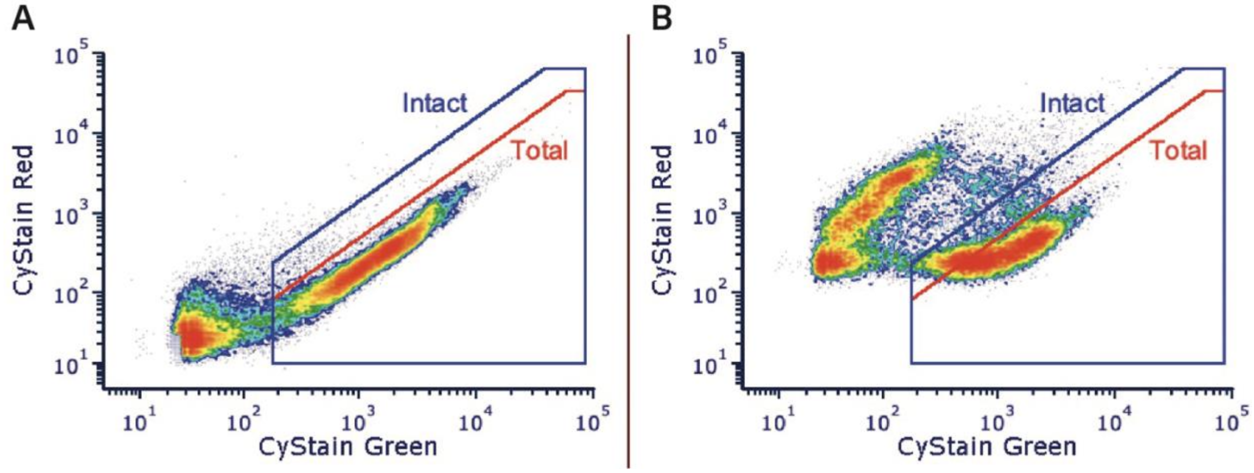


**Figure S2.** Typical flow cytometry event plots, with red fluorescence (CyStain Red) plotted versus green fluorescence (CyStain Green) using the dual-gating strategy. Events within the red "Total" gate in Panel A were counted for total cell counts (TCC). Events within the blue "Intact" gate in Panel B were counted for intact cell counts (ICC). Data shown are from an ozone influent sample taken on Day 29.

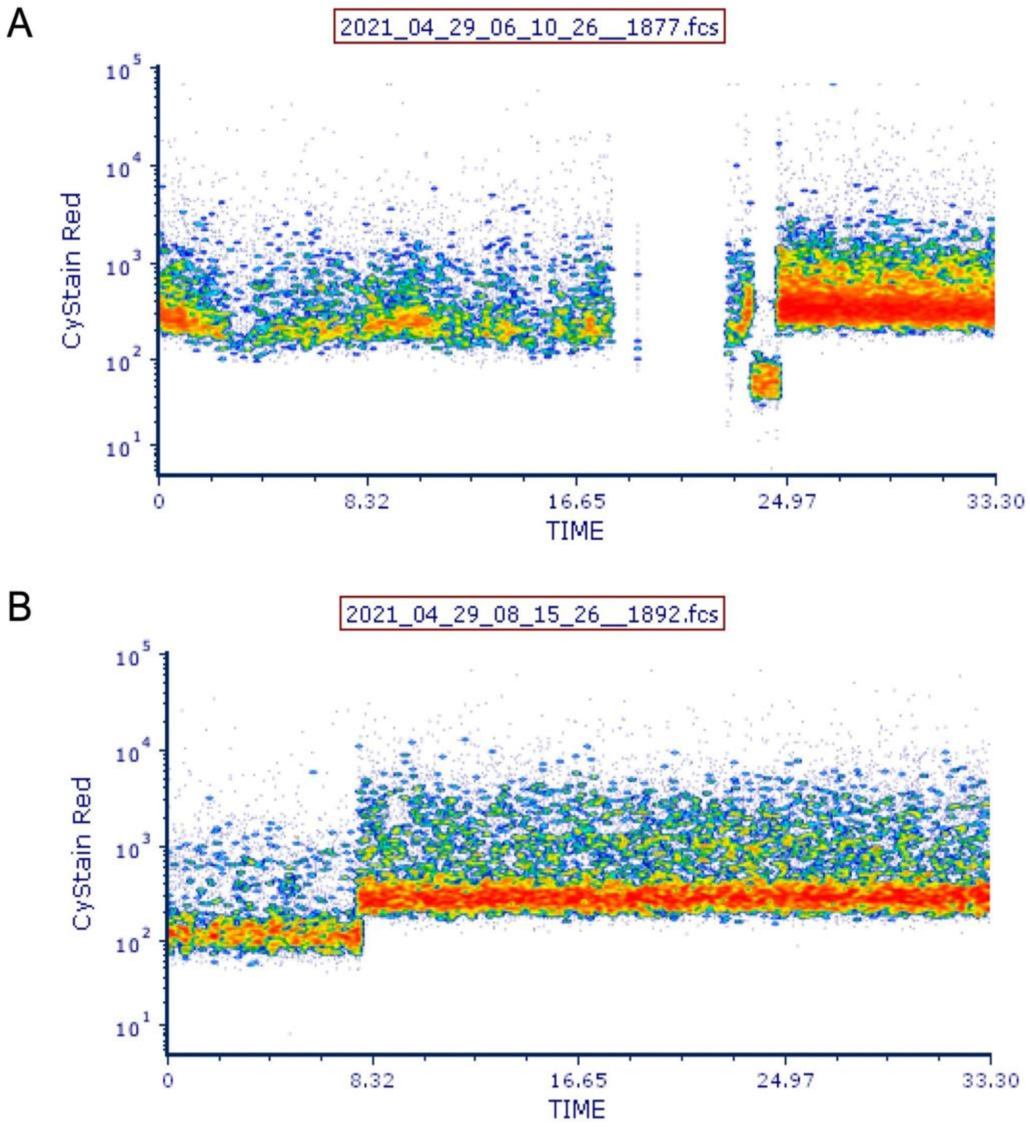


**Figure S3**. Flow cytometry events versus time plots (red fluorescence, “CyStain Red”, versus time in seconds) showing examples of outlier data points resulting from changes in sample flow rate in the flow cytometer during analysis. Panel A shows an example of a sample with a gap in data during acquisition, resulting in periods without few or no events. Panel B shows an example where a sample flow rate change resulted in a jump in fluorescence during the measurement.

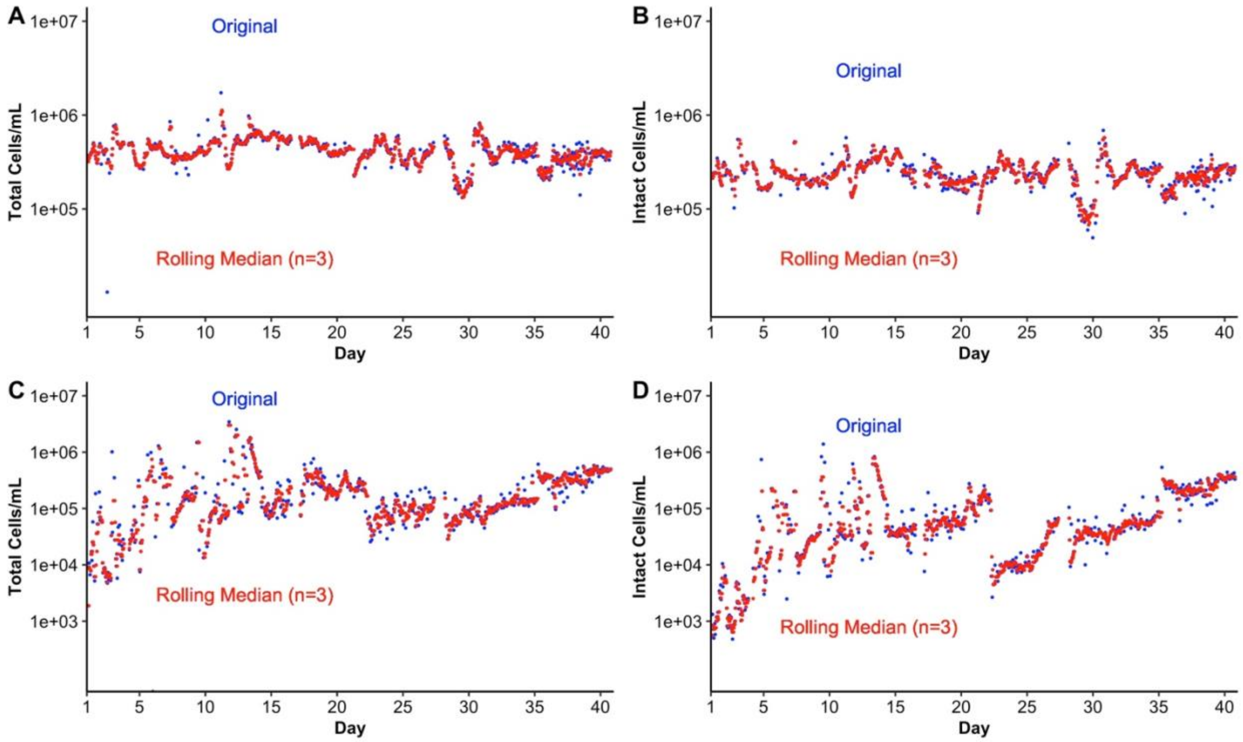


**Figure S4.** Flow cytometry results obtained as described in Figure S2 (blue) and using a rolling median smoothing approach (red), where the median was calculated for three measurements. Plots show the cells per mL versus the date for the A) influent total cell counts (TCC), B) influent intact cell counts (ICC), C) effluent TCC, and D) effluent ICC.

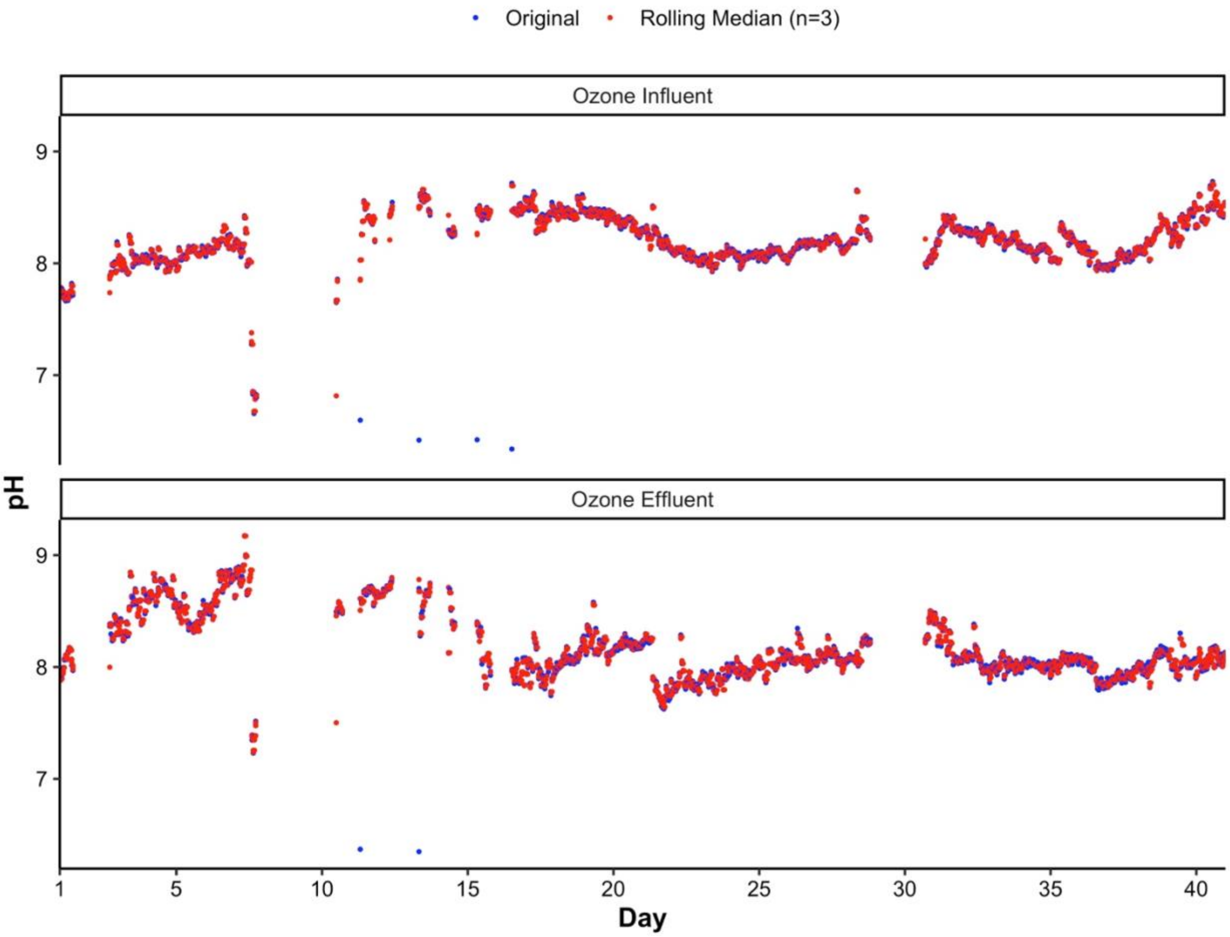


**Figure S5.** Sensor node pH results (blue) and the same data presented using a rolling median smoothing approach (three measurements; red) for the ozone contactor influent and effluent.

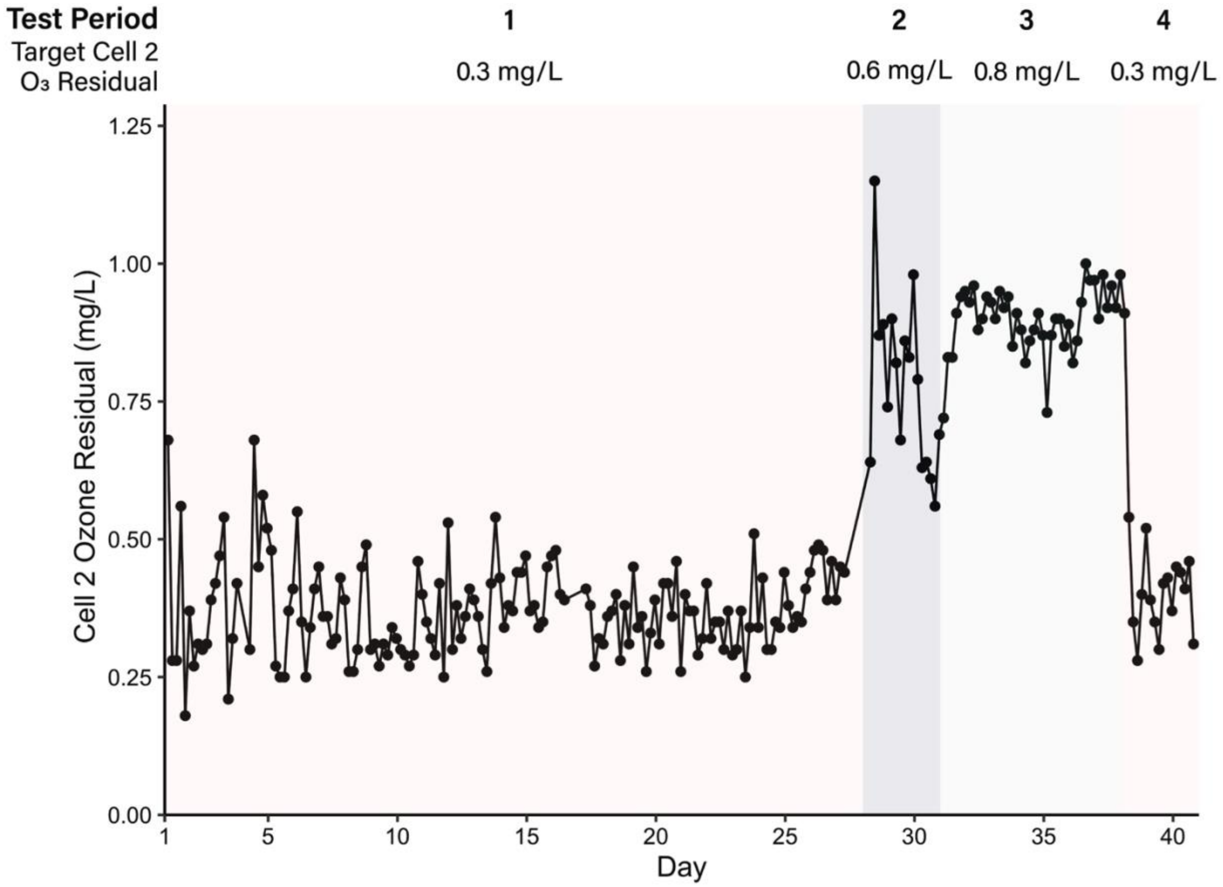


**Figure S6.** Ozone contactor Cell 2 ozone residuals over time during the four test periods. Grab samples collected every four hours. Test Period 1: Target Cell 2 ozone residual of 0.3 mg/L. Test Period 2: Target Cell 2 ozone residual of 0.6 mg/L. Test Period 3: Target Cell 2 ozone residual of 0.8 mg/L. Test Period 4: Target Cell 2 ozone residual of 0.3 mg/L.

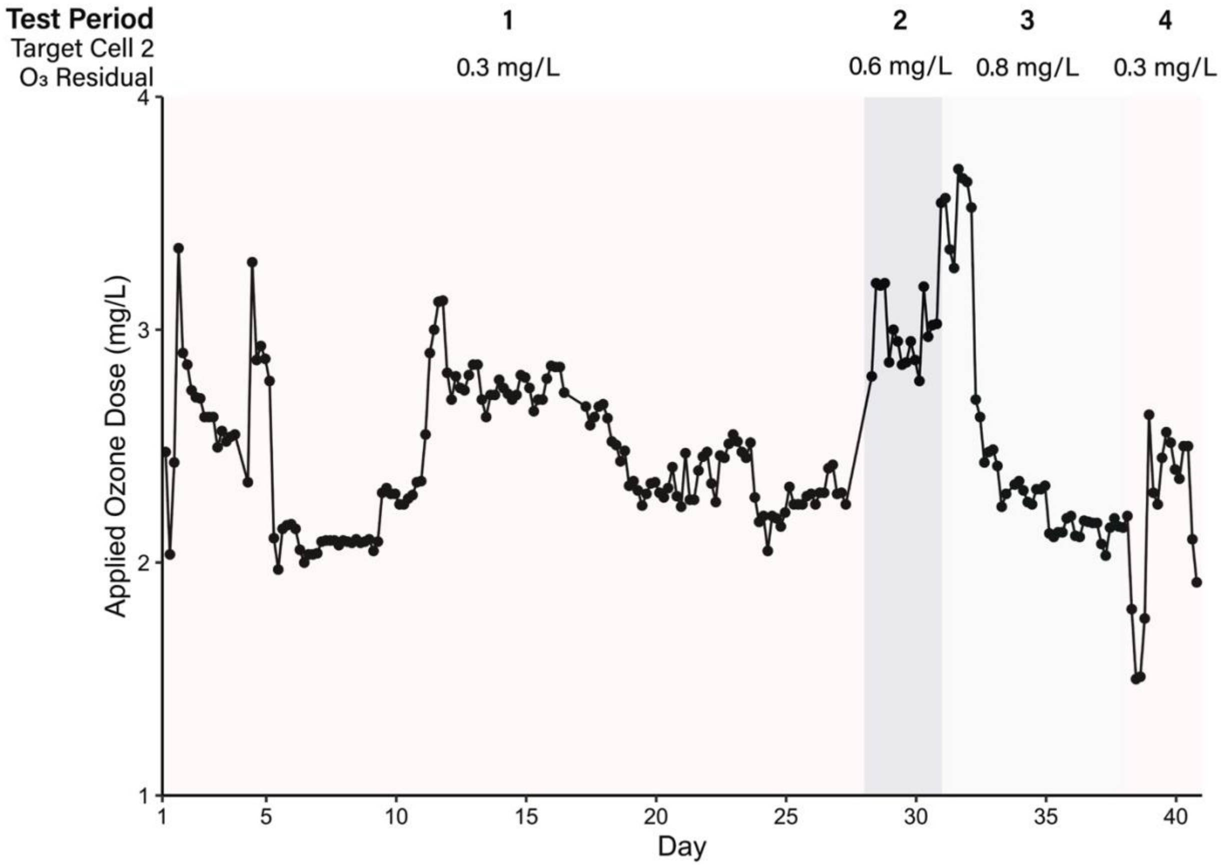


**Figure S7.** Applied ozone dose over time during the four test periods. Calculated from data collected every four hours. Test Period 1: Target Cell 2 ozone residual of 0.3 mg/L. Test Period 2: Target Cell 2 ozone residual of 0.6 mg/L. Test Period 3: Target Cell 2 ozone residual of 0.8 mg/L. Test Period 4: Target Cell 2 ozone residual of 0.3 mg/L.

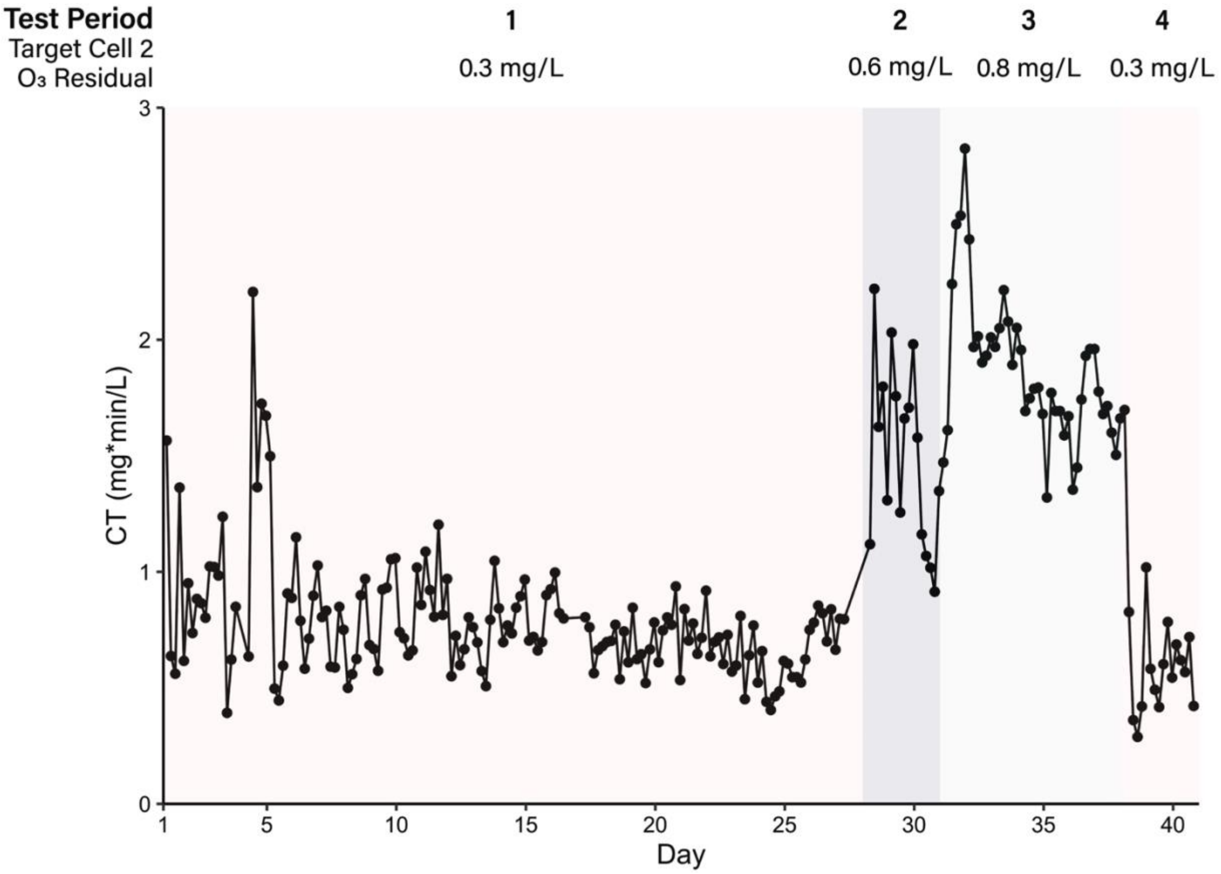


**Figure S8.** Concentration x time (CT) over time during the four test periods. Grab samples collected every four hours. Test Period 1: Target Cell 2 ozone residual of 0.3 mg/L. Test Period 2: Target Cell 2 ozone residual of 0.6 mg/L. Test Period 3: Target Cell 2 ozone residual of 0.8 mg/L. Test Period 4: Target Cell 2 ozone residual of 0.3 mg/L.

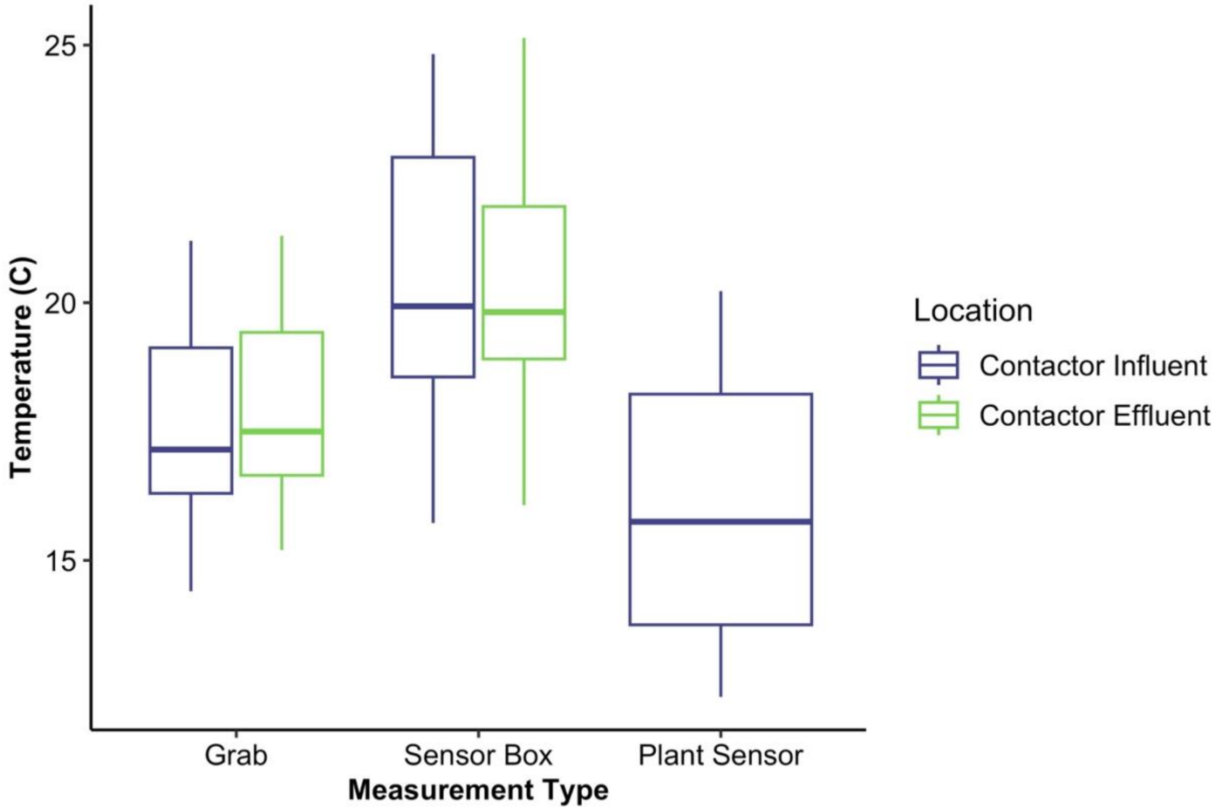


**Figure S9.** Temperature results for the ozone contactor influent and effluent using grab sampling, the sensor boxes, and the water treatment plant's sensor. Sensor box data are represented by the rolling median of three values.

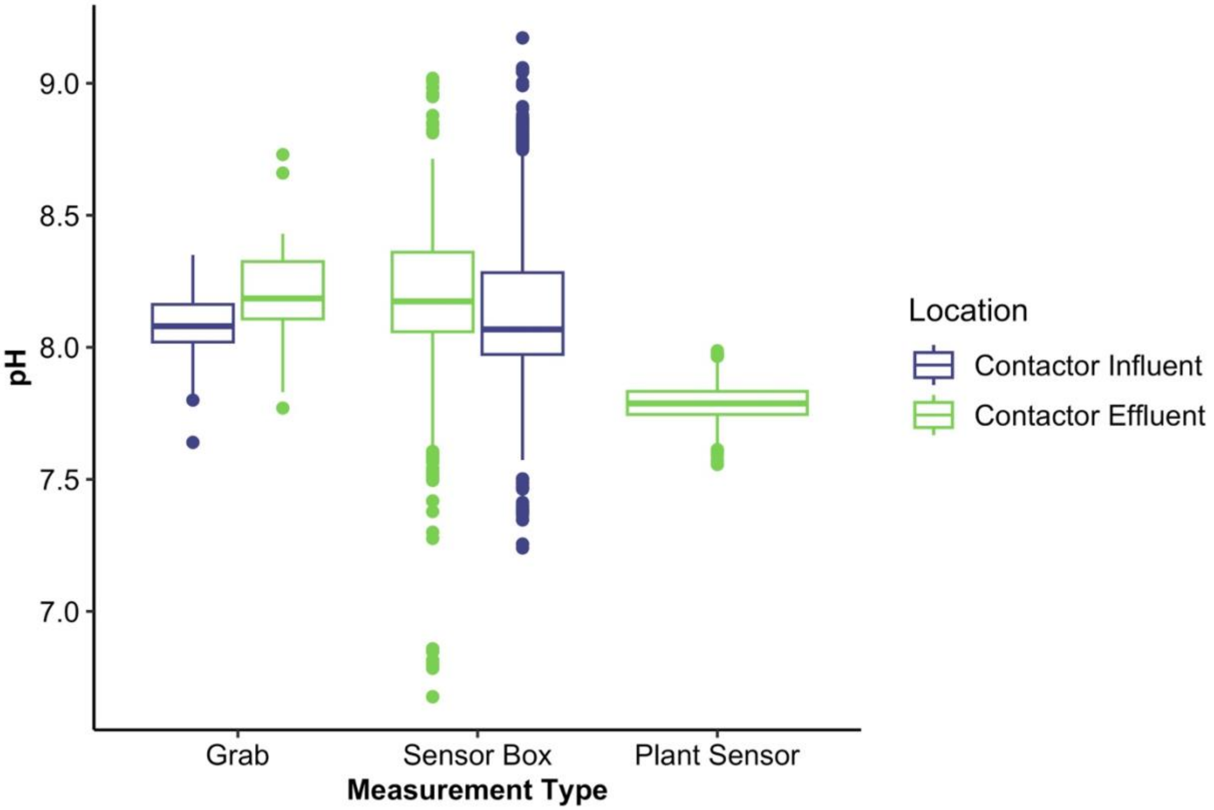


**Figure S10.** pH results for the ozone contactor influent and effluent using grab sampling, the sensor boxes, and the water treatment plant's sensor. Sensor box data are represented by the rolling median of three values.

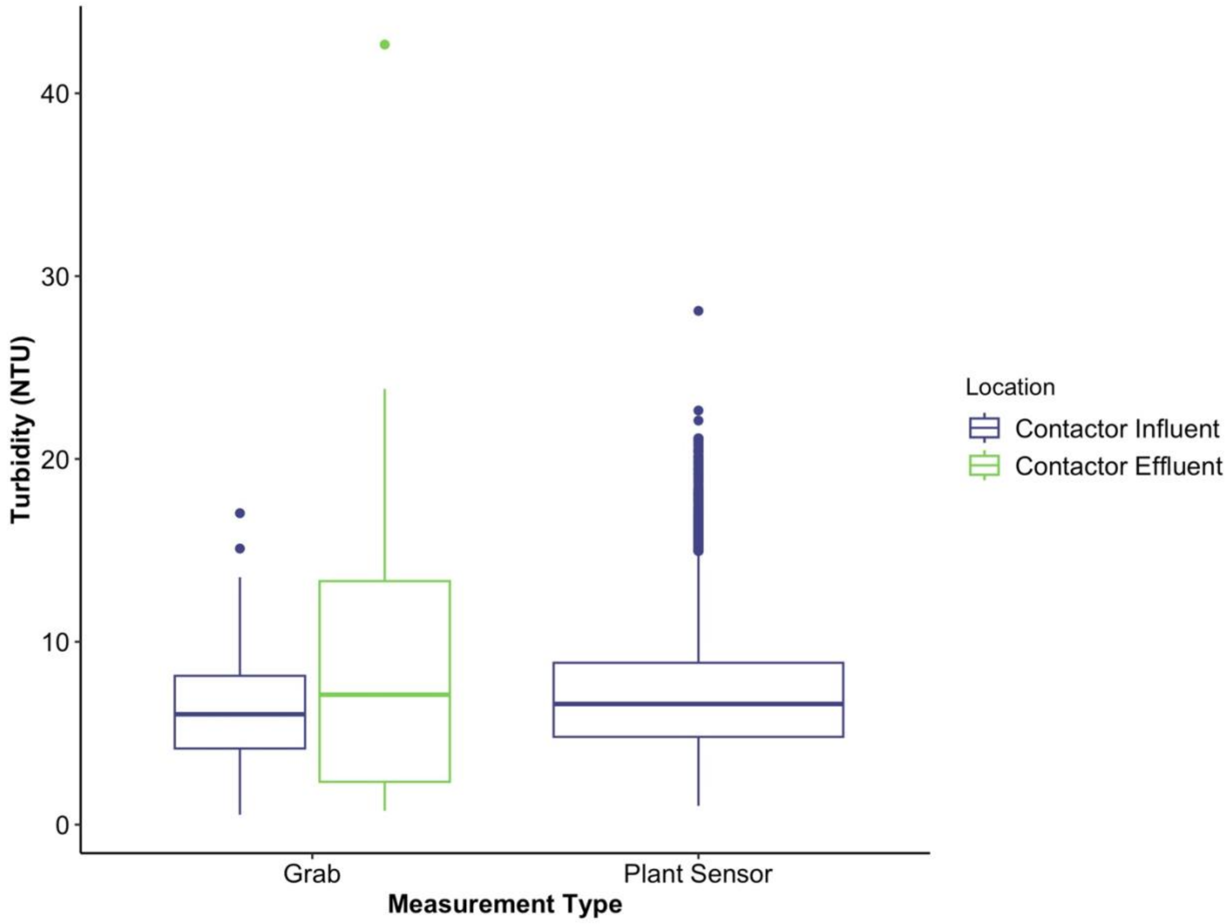


**Figure S11.** Turbidity results for the ozone contactor influent and effluent using grab sampling and the water treatment plant's sensor. Results are in nephelometric turbidity units (NTU).

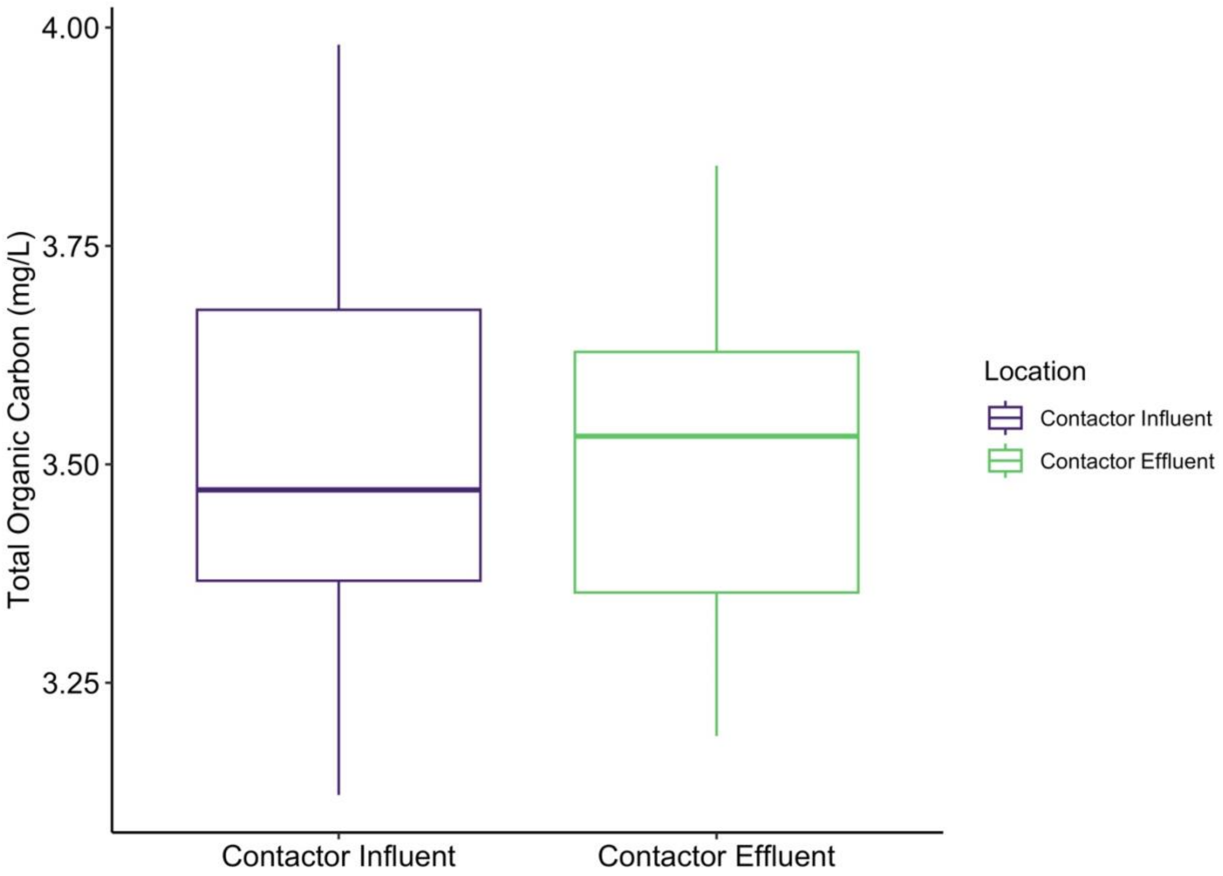


**Figure S12.** Total organic carbon results for the influent and effluent over the study period. Samples were collected in triplicate and six individual readings were done per replicate, yielding 18 measurements per sample.

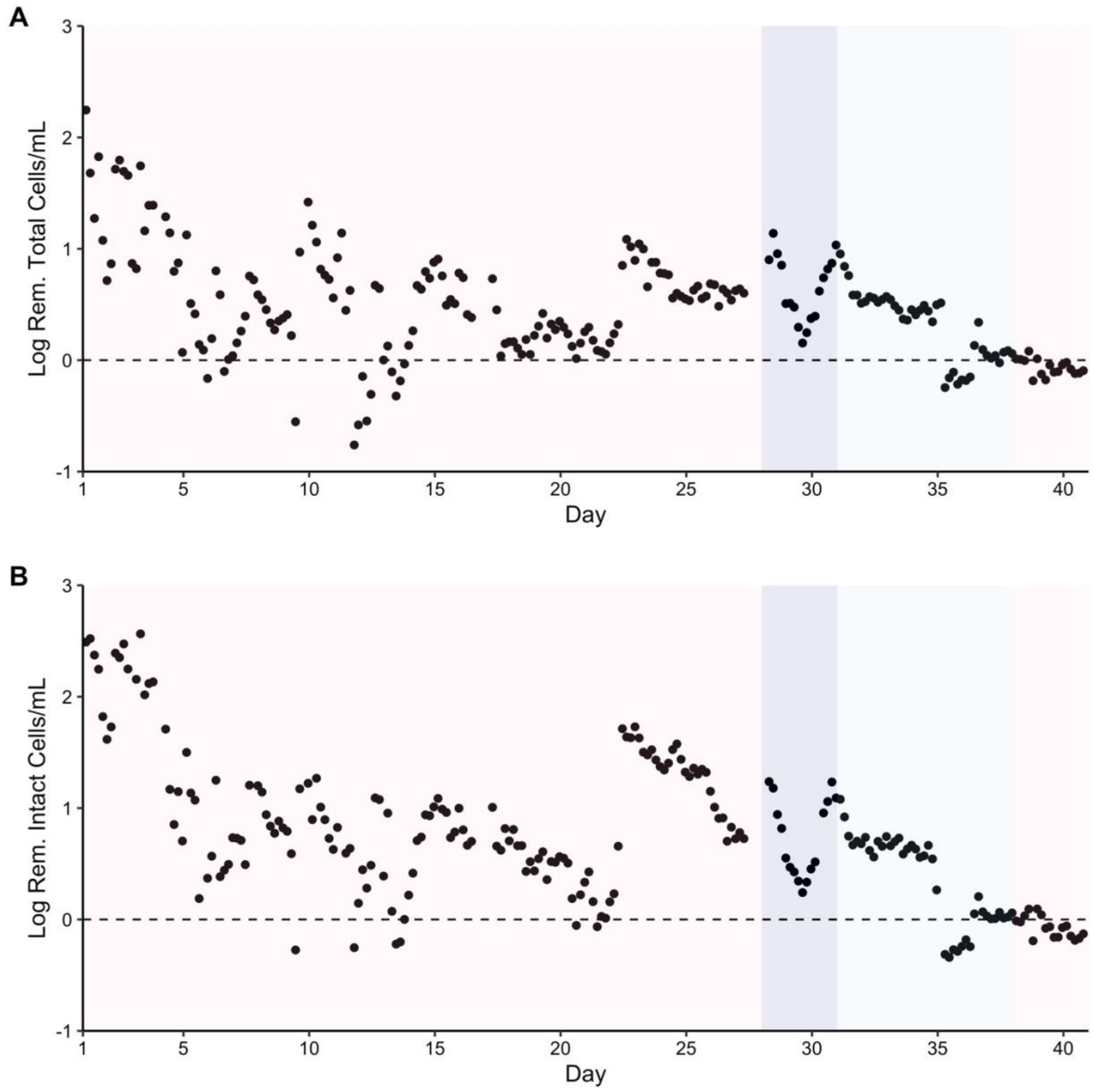


**Figure S13.** A) Total and B) intact cell net log removal over time during the four test periods. Test Period 1: Target Cell 2 ozone residual of 0.3 mg/L. Test Period 2: Target Cell 2 ozone residual of 0.6 mg/L. Test Period 3: Target Cell 2 ozone residual of 0.8 mg/L. Test Period 4: Target Cell 2 ozone residual of 0.3 mg/L.

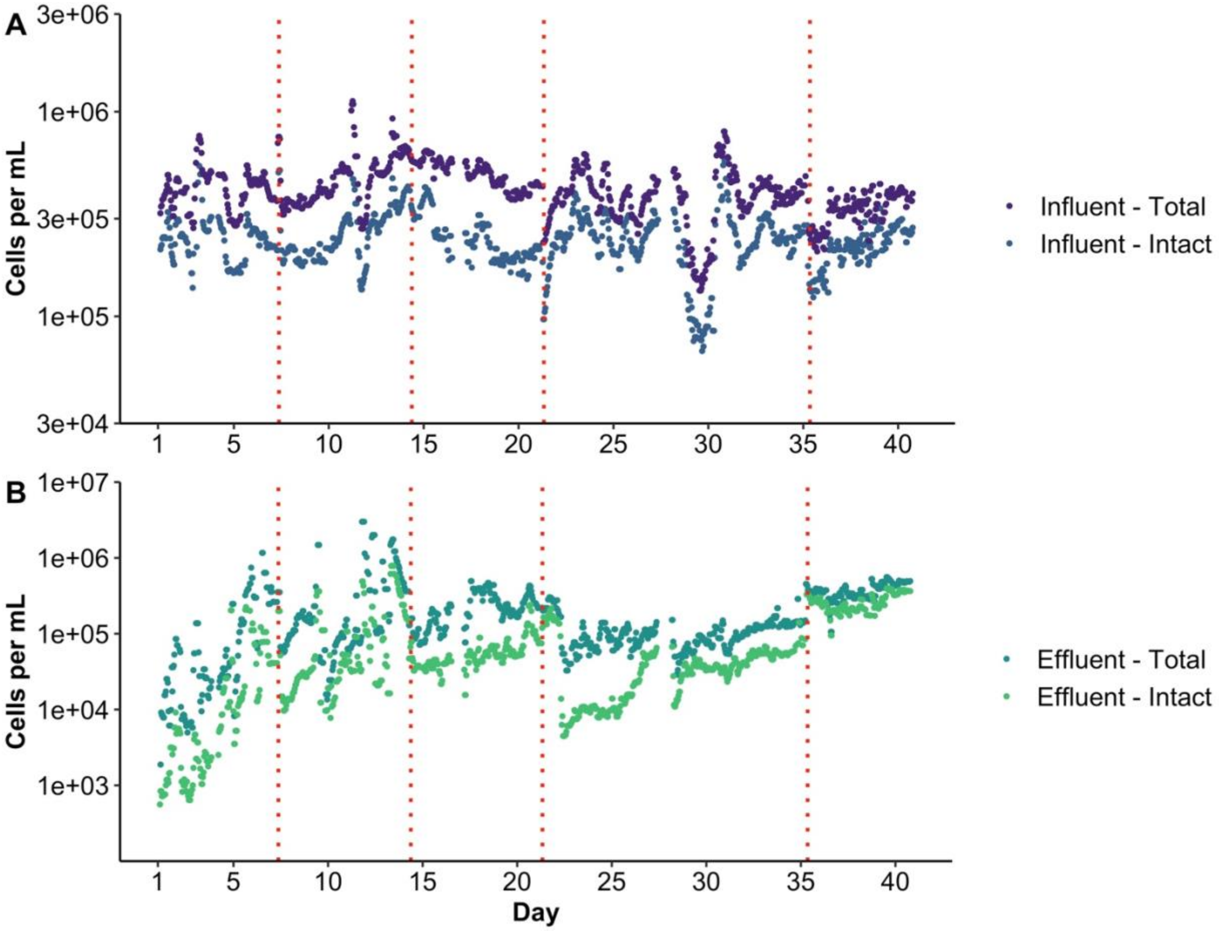


**Figure S14.** Impact of intensive cleaning events on the NRT-FCM results in the A) ozone contactor influent and B) effluent. The vertical red dotted lines indicate intensive cleaning events.

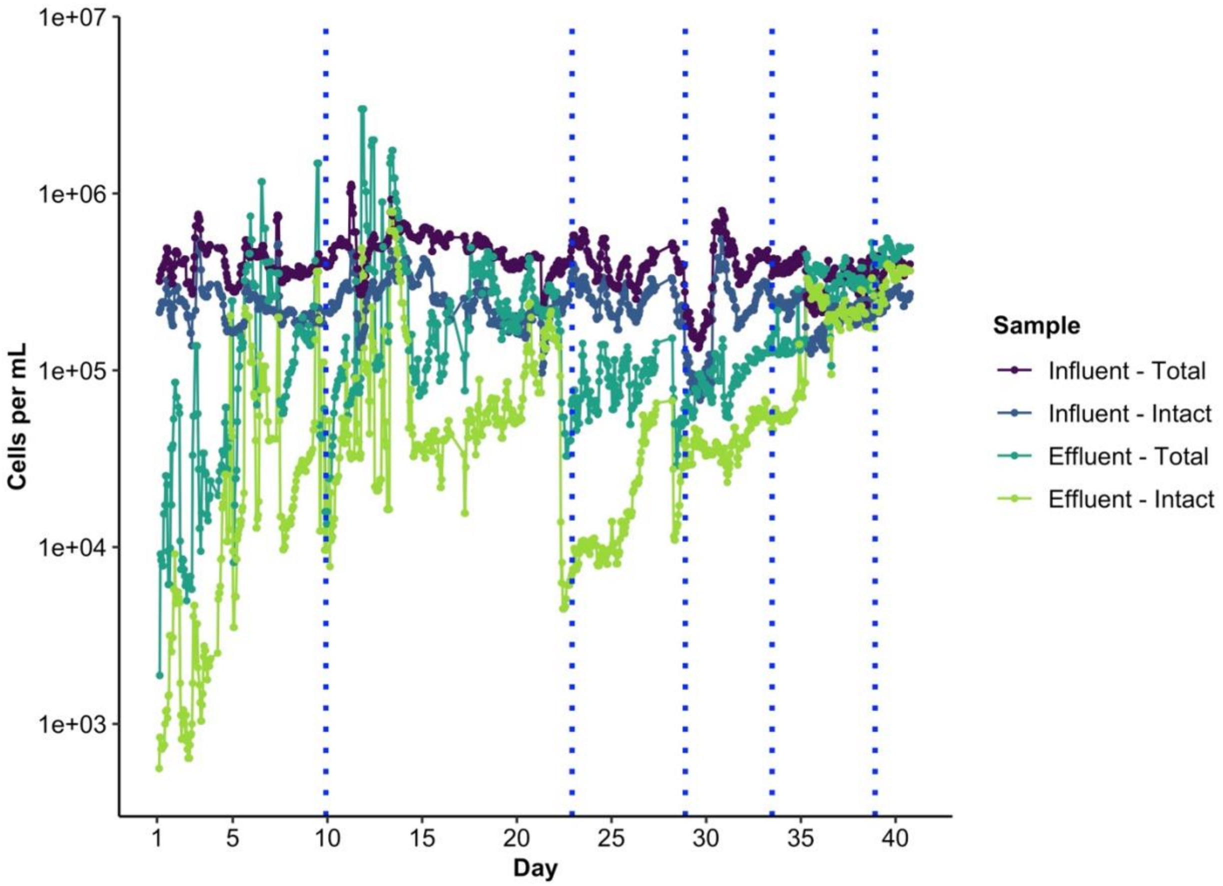


**Figure S15.** Impact of volume deviations during testing on the NRT-FCM results. The blue dotted lines indicate events where it was observed that the volume analyzed exceeded 100 μL.

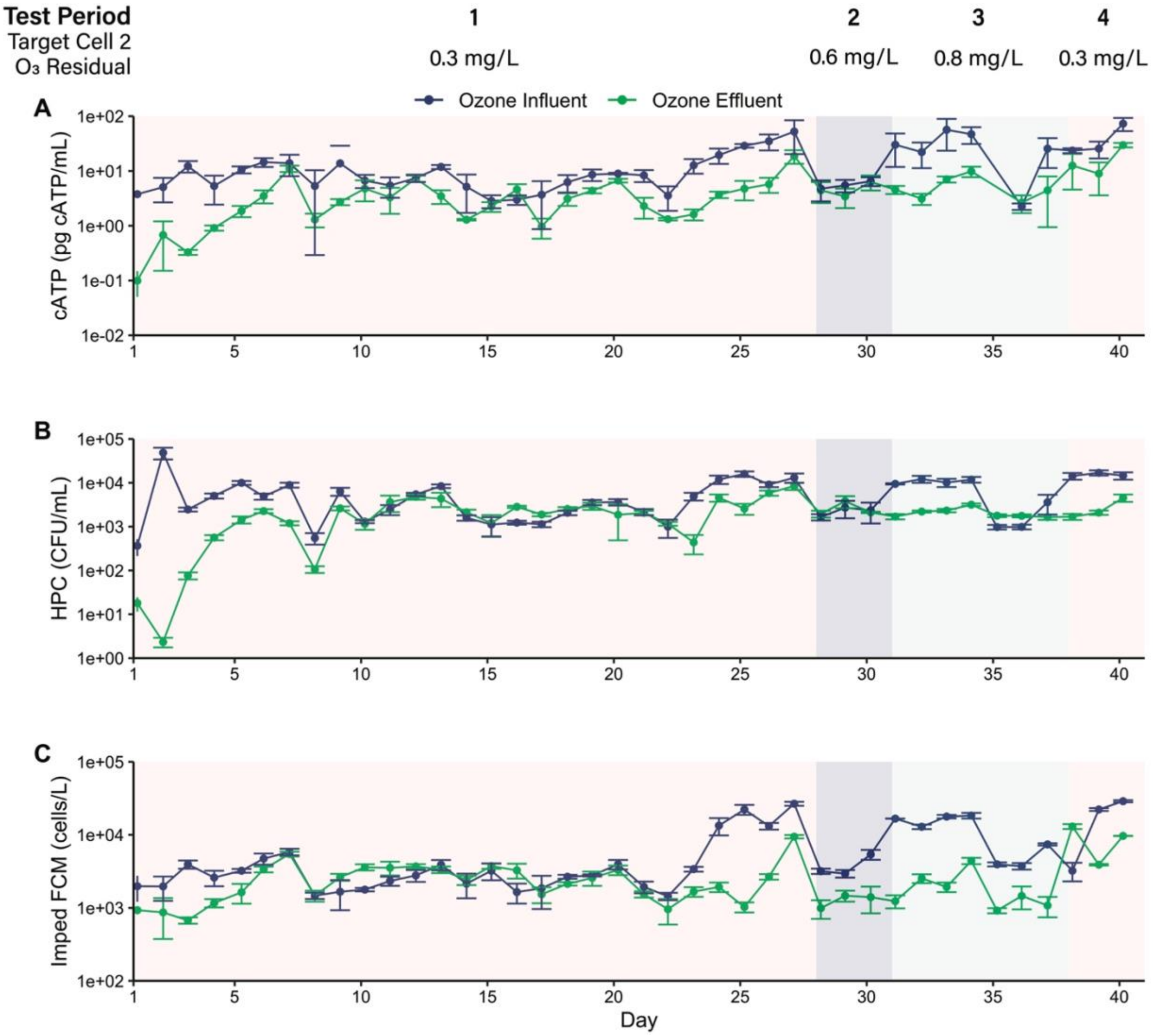


**Figure S16.** Daily microbial grab sample results over time. A) Cellular adenosine triphosphate (cATP), B) heterotrophic plate counts (HPCs), and C) impedance flow cytometry (impedance FCM). Points are the mean of three replicates and error bars show ± one standard deviation.

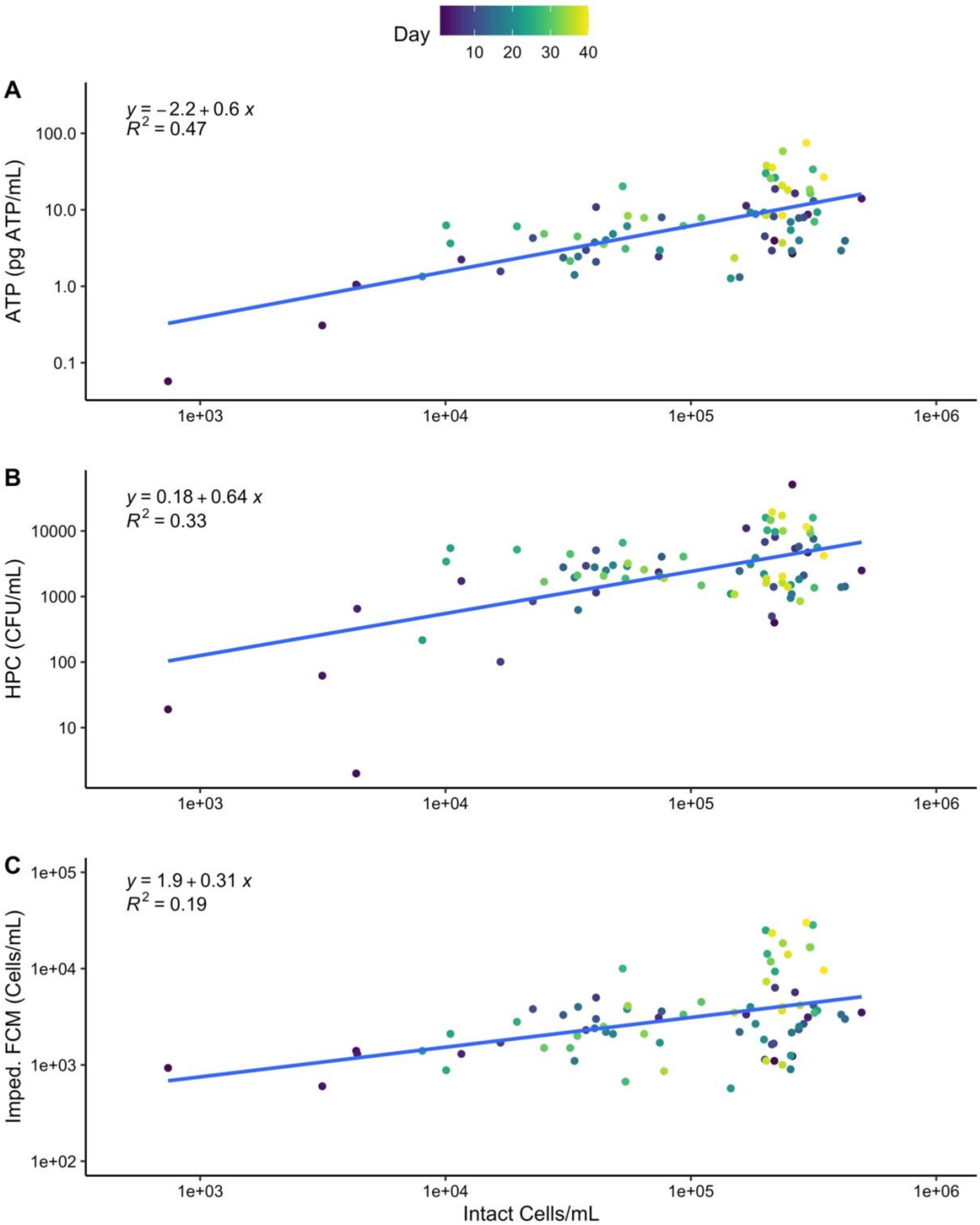


**Figure S17.** Scatter plots of near real-time flow cytometry intact cell counts (ICC) per mL versus A) cellular adenosine triphosphate (cATP), B) heterotrophic plate counts (HPC), and C) impedance flow cytometry (impedance FCM). HPC, cATP, and impedance FCM results are the median of three replicates. Points are colored by the day of the experiment. The blue line shows the linear regression.

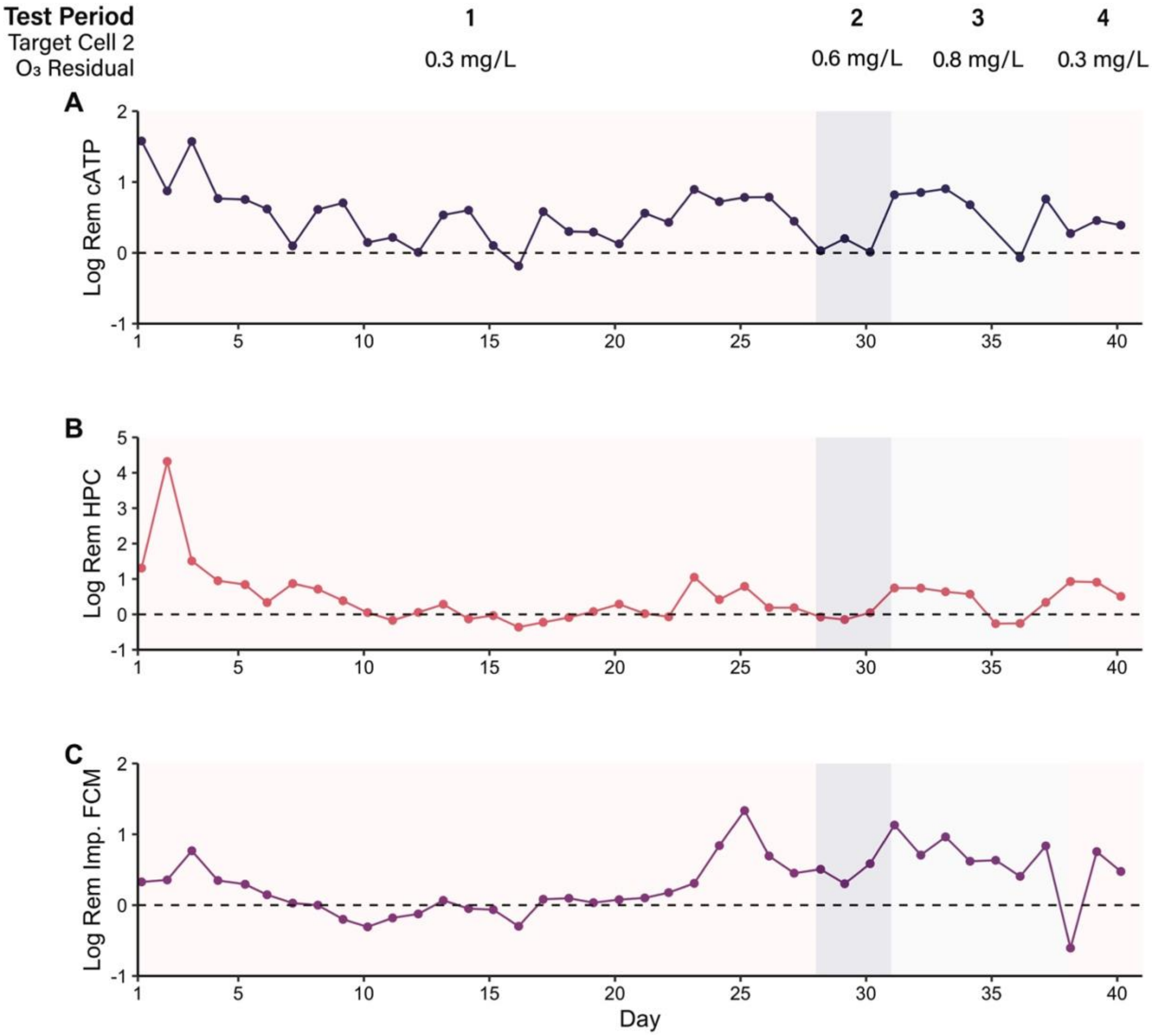


**Figure S18.** Net log removal over time for the daily microbial grab samples. A) Cellular adenosine triphosphate (cATP), B) heterotrophic plate counts (HPC), and C) impedance flow cytometry (Imp. FCM). The dashed line indicates zero log removal.

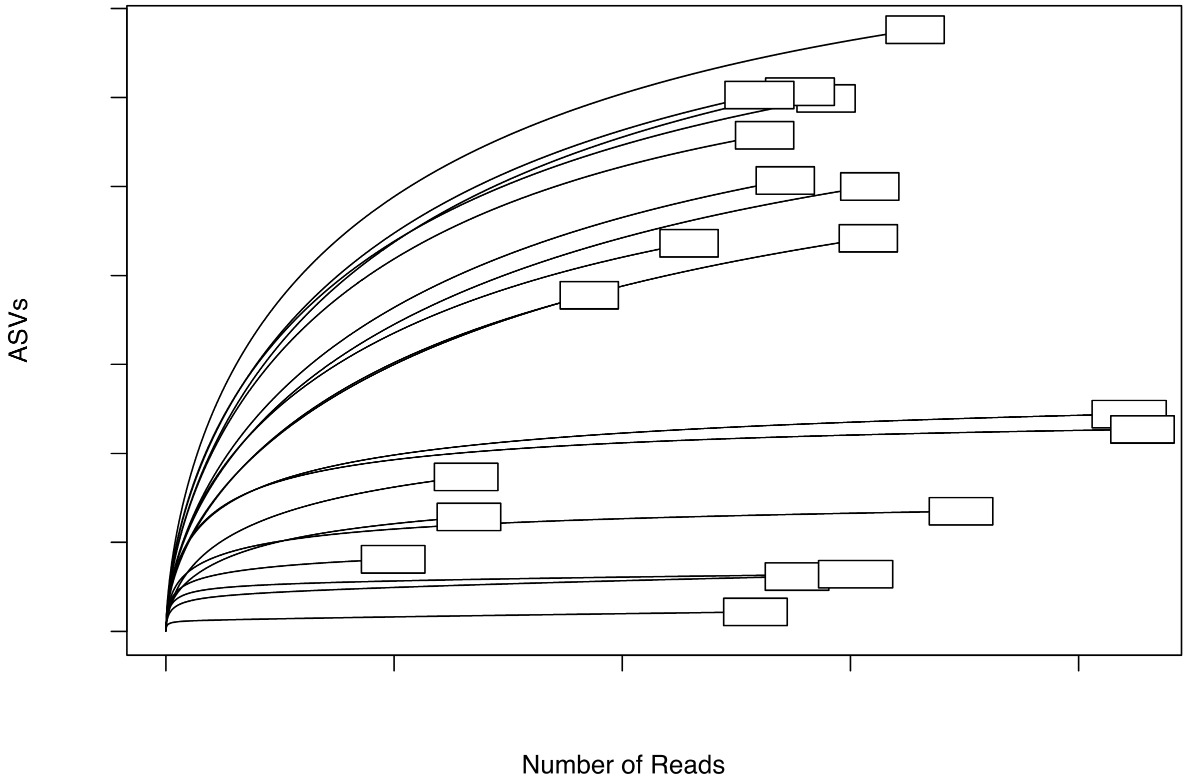


**Figure S19.** Rarefaction curves for 16S rRNA gene sequencing results showing the number of amplicon sequence variants (ASVs) versus the number of reads for each sample. INF: Ozone contactor influent. EFF: Ozone contactor effluent. The number in the sample name indicates the sampling event (1-11). Two ozone effluent samples (EFF-4 and EFF-6) were excluded from analyses due to low read counts (less than 1,000 reads). One influent sample (INF-6) was excluded due to contamination.

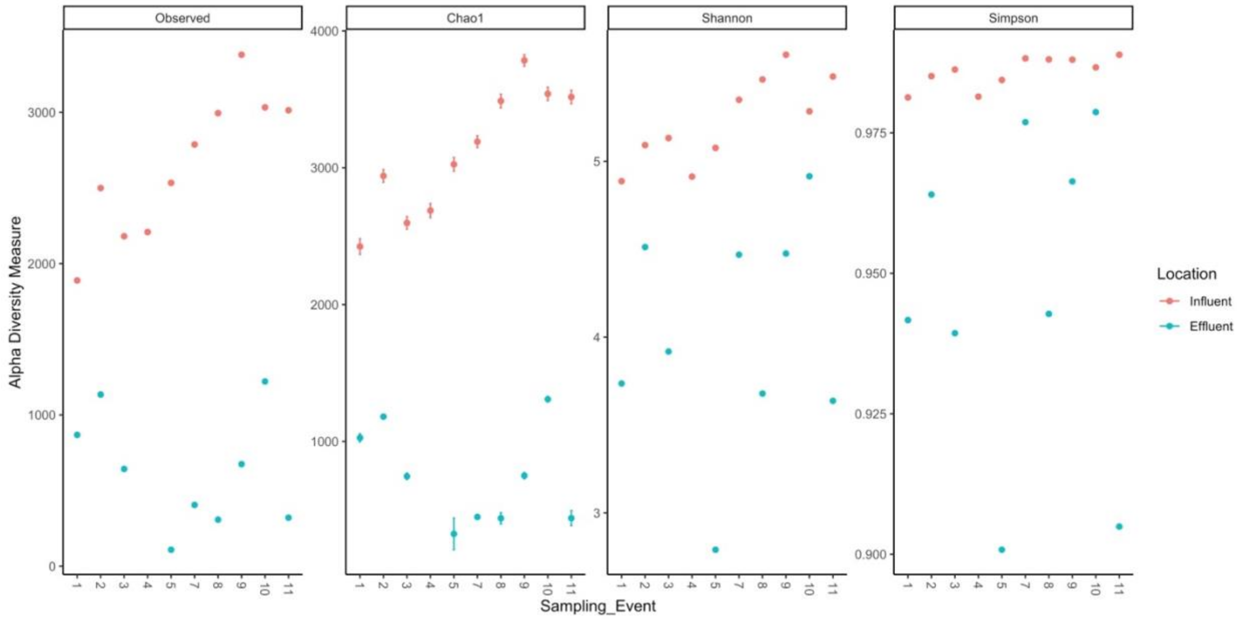


**Figure S20.** Alpha diversity results (observed amplicon sequence variants [ASVs], Chao1, Shannon, and Simpson) for the ozone contactor influent and effluent across the 11 sampling events. Ozone contactor influent samples ("Influent") are in red and ozone contactor effluent samples ("Effluent") are in blue. Sampling event dates: 1 – Day 3 (April 30, 2021); 2 – Day 7 ( May 4, 2021); 3 – Day 10 (May 7, 2021); 4 – Day 14 (May 11, 2021); 5 – Day 17 (May 14, 2021); 6 – Day 21 (May 18, 2021); 7 – Day 24 (May 21, 2021); 8 – Day 28 (May 25, 2021); 9 – Day 30 (May 27, 2021); 10 – Day 35 (June 1, 2021); 11 – Day 38 (June 4, 2021). Two ozone effluent samples (EFF-4 and EFF-6) were excluded from analyses due to low read counts (less than 1,000 reads). One influent sample (INF-6) was excluded due to contamination.

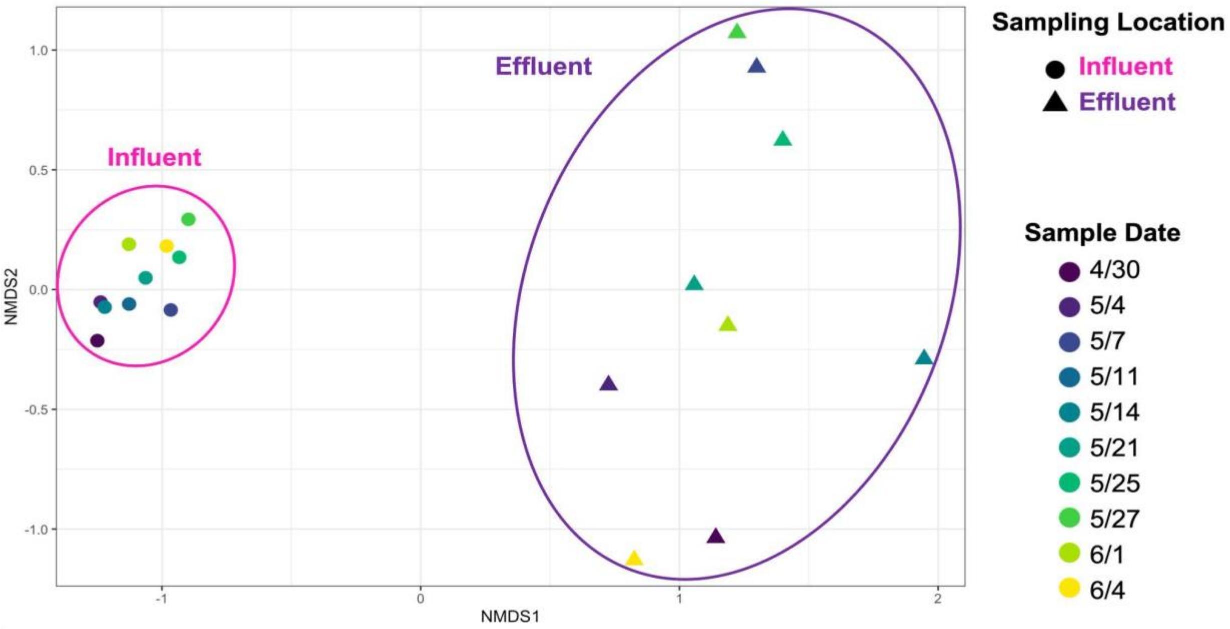


**Figure S21.** Results of non-metric multidimensional scaling (NMDS) analysis for the 16S rRNA gene sequencing results. The ozone contactor influent ("Influent") samples are shown as circles and the ozone contactor effluent ("Effluent") samples are shown as triangles. Points are colored by the sampling date, ranging from dark purple in late April and early May 2021 to yellow in early June 2021.

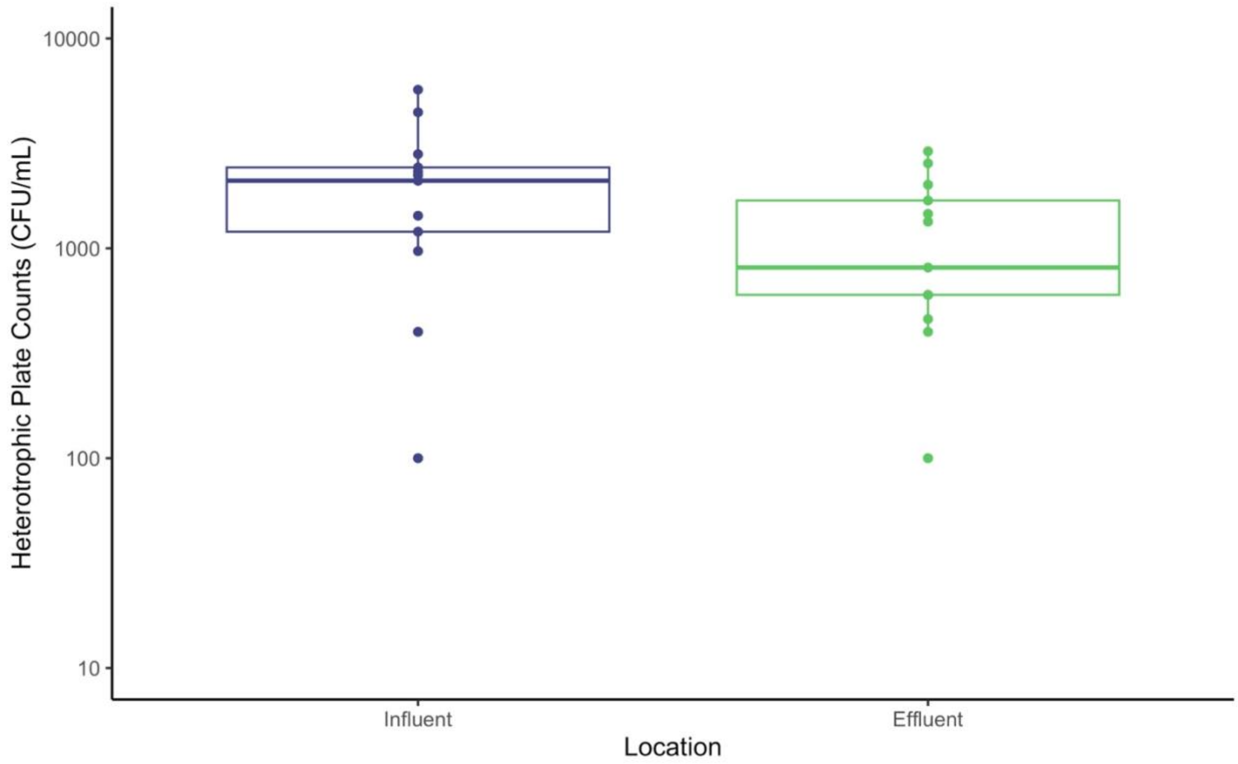


**Figure S22.** Heterotrophic plate count (HPC) results during 2022 follow-up testing. Ozone contactor influent: “Influent”. Ozone contactor effluent: “Effluent”.

#### Supplementary Equations

**Equation S1. T10 calculation**

$$T10\ (min) = cell\ vol\ (cubic\ feet) \times \frac{\#\ contactors\ in\ service}{plant\ flow\ (MGD)} \times \frac{1440\ min}{1\ day} \times \frac{1\ million\ gal}{10^6\ gal} \times \frac{7.48\ gal}{cubic\ foot} \times baffle\ factor$$

**Equation S2. Cell 2 average ozone residual and CT calculations**

$$Cell\ 2\ average\ ozone\ residual\ \left(Cell\ 2_{avg}, \frac{mg}{L}\right) = \frac{Cell\ 2\ effluent\ ozone\ residual\ (Cell\ 2_{eff\ resid}, \frac{mg}{L})}{2}$$

- If $Cell\ 2_{avg} < 0.1 \frac{\text{mg}}{\text{L}}, Cell\ 2\ CT = 0 \frac{\text{mg*min}}{\text{L}}$
- If $Cell\ 2_{avg} \geq 0.1 \frac{mg}{L}, Cell\ 2\ CT = T10 \times\ Cell\ 2_{avg}$

**Equation S3. Cell 3 average ozone residual and CT calculations**

$$Cell\ 3\ average\ ozone\ residual\ (Cell\ 3_{avg}) = \frac{Cell\ 2_{eff\ resid}\left(\frac{mg}{L}\right) +\ Cell\ 3\ effluent\ ozone\ residual\ (Cell\ 3_{eff\ resid}, \frac{mg}{L})}{2}$$

- If $Cell\ 3_{avg} < 0.1 \frac{mg}{L}, Cell\ 3\ CT = 0 \frac{\text{mg*min}}{\text{L}}$
- If $Cell\ 3_{avg} \geq 0.1 \frac{mg}{L}, Cell\ 3\ CT = T10 \times\ Cell\ 3_{avg}$

**Equation S4. Cell 4 CT calculation**

$$Cell\ 4\ CT = T10\ \times\ Cell\ 4\ effluent\ ozone\ residual$$

**Equation S5.** Applied ozone dose calculation

$$Applied\ O_3\ dose\ \left(\frac{mg}{L}\right) = \frac{fraction\ O_3\ \left(\frac{\%\ O_3\ gas}{100}\right) \times gas\ flow \left(\frac{lbs}{day}\right) \times 453{,}592\,\frac{mg}{lb}}{plant\ flow\ \left(\frac{10^6\ gal}{day}\right) \times \left(\frac{3.785\ L}{gal}\right)}$$

**Equation S6.** Conversion from relative light units (RLU) to cellular ATP (cATP) per mL of sample

$$cATP\left(\frac{pg\ ATP}{mL}\right)=\frac{RLU_{cATP}}{RLU_{ATP1}}\times\frac{10{,}000\ (pg\ ATP)}{V_{sample}(mL)}$$

Where

$RLU_{cATP}$ is the result from the sample

$RLU_{ATP}$ is the result for the Luminase

### Supplementary Tables

**Table S1.** Sample Analysis Methods

| Category | Analysis | Sample Locations | Instrument | Method | Detection Limit(s)[1] | Sample Type | Frequency | Replication | Collection Vessel | | |
|---|---|---|---|---|---|---|---|---|---|---|---|
| | | | | | | | | | Description | Manufacturer | Cat No. |
| Physico-chemical | Electrical Conductivity | Inf & Eff | Atlas Scientific #ENV-40-EC-K1.0 | Graphite measuring surface with K=1 | 5 µS/cm – 200,000 µS/cm | Continuous | hourly | triplicate | N/A | | |
| | Oxidation-Reduction Potential (ORP) | Inf & Eff | Atlas Scientific #ENV-40-ORP | Reference electrode silver/silver chloride. 4M KCl reference solution, platinum tip | (+/-) 2,000 mV | Continuous | hourly | triplicate | N/A | | |
| | Ozone Residual | Cells 2, 3, 4 | Hach | Method 8311 | 0.01 mg/L | Grab | every 4 hrs | N/A | plastic beaker | | |
| | | Cell 2 | Rosemount™ 56 Dual Channel Transmitter with dissolved ozone (Rosemount 499AOZ) and temperature sensors | | LLOD: 0.001 ULOD: 1 mg/L | Continuous | every 5 mins | N/A | | | |
| | pH | Inf & Eff | Not provided | | --- | Grab | daily | N/A | plastic beaker | | |
| | | Inf & Eff | Atlas Scientific #ENV-40-pH | Silver/silver chloride with KCl reference solution and EXR advanced sensing glass | --- | Continuous | hourly | triplicate | N/A | | |
| | | Influent | Not provided | | --- | Continuous | every 5 mins | N/A | | | |
| | Temperature | Inf & Eff | Not provided | | --- | Grab | daily | N/A | plastic beaker | | |
| | | Inf & Eff | Atlas Scientific #PT-1000 | Thermocouple | --- | Continuous | hourly | triplicate | N/A | | |
| | | Influent | Not provided | | --- | Continuous | every 5 mins | N/A | | | |
| | Total Organic Carbon (TOC) | Inf & Eff | Shimadzu TOC-V | EPA 415.3 | 0.1 mg/L | Grab | daily | triplicate | Carbon-free 40 mL amber glass vials | Fisher Scientific | 12100108 |
| | Turbidity | Inf & Eff | Hach TU5200 | SM 2130 B-01 | 0.01 NTU | Grab | daily | triplicate | Hach glass measurement vial | | |
| | | Effluent | Hach 1720 C | SM 2130 B | 0.003 NTU | Continuous | every 5 mins | N/A | | | |

| | | | | | | | | | | | |
|---|---|---|---|---|---|---|---|---|---|---|---|
| Biological | Adenosine Triphosphate | Inf & Eff | LuminUltra PhotonMaster Luminometer | Quench-Gone Aqueous, ASTM Std D4012 | <10 relative light units (RLU) | Grab | daily | triplicate | 120 mL sterile sample containers with sodium thiosulfate | IDEXX Laboratories Inc. | WV120SBST-20 |
| | DNA | Inf & Eff | N/A | Collection onto 0.2-micron Sterivex filters | --- | Grab | 2x per week | N/A | sterile 10 L carboy | | |
| | Heterotrophic Plate Counts | Inf & Eff | N/A | SM 9215-B-2000 | <1 CFU/ vol | Grab | daily | triplicate | 120 mL sterile sample containers with sodium thiosulfate | IDEXX Laboratories Inc. | WV120SBST-20 |
| | Intact Cell Counts | Inf & Eff | BactoBox | Impedance Flow Cytometry | LLOD: 100 ULOD: 2,000 cells/mL | Grab | daily | triplicate | 120 mL sterile sample containers with sodium thiosulfate | IDEXX Laboratories Inc. | WV120SBST -20 |
| | Total and Intact Cell Counts | Inf & Eff | Sysmex | Flow Cytometry | 1 cell per 100 µL | Continuous | every ~20 mins | N/A | | | |

1. Lower limit of detection (LLOD) unless otherwise specified. ULOD: Upper limit of detection.

**Table S2.** Filtered volumes for DNA analyses

| Sampling Event | Sampling Date | Experiment Day | Sample Volume (L) | |
|---|---|---|---|---|
| | | | Ozone Contactor Influent | Ozone Contactor Effluent |
| 1 | April 30, 2021 | 3 | 3.2 | 5.8 |
| 2 | May 4, 2021 | 7 | 5.2 | 8.3 |
| 3 | May 7, 2021 | 10 | 6.6 | 9.0 |
| 4 | May 11, 2021 | 14 | 4.5 | 7.8 |
| 5 | May 14, 2021 | 17 | 4.2 | 8.3 |
| 6 | May 18, 2021 | 21 | 4.7 | 8.1 |
| 7 | May 21, 2021 | 24 | 5.6 | 6.2 |
| 8 | May 25, 2021 | 28 | 6.0 | 9.6 |
| 9 | May 27, 2021 | 30 | 9.0 | 10.5 |
| 10 | June 1, 2021 | 35 | 4.9 | 6.4 |
| 11 | June 4, 2021 | 38 | 7.2 | 9.4 |

**Table S3.** 16S rRNA sequencing top 30 ASVs and relative abundances as a fraction of one

| ASV | Kingdom | Phylum | Class | Order | Family | Genus | Species | INF-1 | EFF-1 | INF-2 | EFF-2 | INF-3 | EFF-3 | INF-4 | INF-5 | EFF-5 | INF-7 | EFF-7 | INF-8 | EFF-8 | INF-9 | EFF-9 | INF-10 | EFF-10 | INF-11 | EFF-11 |
|---|---|---|---|---|---|---|---|---|---|---|---|---|---|---|---|---|---|---|---|---|---|---|---|---|---|---|
| ASV10 | Bacteria | Actinobacteriota | Actinobacteria | Frankiales | Sporichthyaceae | NA | NA | 0.079 | 0.010 | 0.074 | 0.016 | 0.057 | 0.001 | 0.060 | 0.078 | 0.000 | 0.087 | 0.018 | 0.063 | 0.003 | 0.053 | 0.001 | 0.070 | 0.003 | 0.085 | 0.010 |
| ASV11 | Bacteria | Bacteroidota | Bacteroidia | Cytophagales | Spirosomaceae | *Pseudarcicella* | NA | 0.119 | 0.008 | 0.069 | 0.006 | 0.086 | 0.001 | 0.118 | 0.101 | 0.000 | 0.047 | 0.009 | 0.028 | 0.004 | 0.015 | 0.000 | 0.078 | 0.002 | 0.063 | 0.000 |
| ASV12 | Bacteria | Bacteroidota | Bacteroidia | Chitinophagales | Chitinophagaceae | *Sediminibacterium* | NA | 0.125 | 0.006 | 0.124 | 0.012 | 0.085 | 0.000 | 0.106 | 0.092 | 0.000 | 0.034 | 0.000 | 0.014 | 0.000 | 0.008 | 0.000 | 0.027 | 0.001 | 0.028 | 0.000 |
| ASV13 | Bacteria | Proteobacteria | Gammaproteobacteria | Burkholderiales | Oxalobacteraceae | *Actimicrobium* | NA | 0.000 | 0.017 | 0.000 | 0.014 | 0.000 | 0.147 | 0.000 | 0.000 | 0.020 | 0.000 | 0.032 | 0.000 | 0.250 | 0.000 | 0.032 | 0.000 | 0.010 | 0.000 | 0.001 |
| ASV14 | Bacteria | Bacteroidota | Bacteroidia | Chitinophagales | Chitinophagaceae | *Sediminibacterium* | NA | 0.002 | 0.001 | 0.007 | 0.004 | 0.076 | 0.021 | 0.055 | 0.014 | 0.000 | 0.089 | 0.019 | 0.061 | 0.000 | 0.057 | 0.042 | 0.020 | 0.030 | 0.095 | 0.001 |
| ASV15 | Bacteria | Proteobacteria | Gammaproteobacteria | Burkholderiales | Methylophilaceae | *Candidatus Methylopumilus* | NA | 0.039 | 0.001 | 0.063 | 0.004 | 0.042 | 0.000 | 0.049 | 0.064 | 0.000 | 0.067 | 0.001 | 0.055 | 0.002 | 0.063 | 0.000 | 0.040 | 0.001 | 0.042 | 0.009 |
| ASV16 | Bacteria | Bacteroidota | Bacteroidia | Sphingobacteriales | NS11-12 marine group | NA | NA | 0.031 | 0.000 | 0.045 | 0.000 | 0.048 | 0.000 | 0.039 | 0.056 | 0.000 | 0.064 | 0.002 | 0.034 | 0.000 | 0.052 | 0.000 | 0.071 | 0.001 | 0.133 | 0.000 |
| ASV17 | Bacteria | Proteobacteria | Gammaproteobacteria | Burkholderiales | Comamonadaceae | *Ottowia* | NA | 0.000 | 0.000 | 0.000 | 0.000 | 0.000 | 0.000 | 0.000 | 0.000 | 0.181 | 0.000 | 0.000 | 0.000 | 0.000 | 0.000 | 0.000 | 0.000 | 0.000 | 0.000 | 0.000 |
| ASV18 | Bacteria | Proteobacteria | Gammaproteobacteria | Xanthomonadales | Xanthomonadaceae | *Silanimonas* | NA | 0.003 | 0.000 | 0.012 | 0.000 | 0.011 | 0.000 | 0.008 | 0.016 | 0.000 | 0.033 | 0.000 | 0.127 | 0.000 | 0.178 | 0.000 | 0.131 | 0.002 | 0.048 | 0.000 |
| ASV19 | Bacteria | Cyanobacteria | Cyanobacteriia | Chloroplast | NA | NA | NA | 0.002 | 0.169 | 0.001 | 0.128 | 0.001 | 0.005 | 0.000 | 0.000 | 0.067 | 0.000 | 0.013 | 0.000 | 0.001 | 0.000 | 0.000 | 0.000 | 0.007 | 0.000 | 0.005 |
| ASV2 | Bacteria | Proteobacteria | Gammaproteobacteria | Burkholderiales | Comamonadaceae | *Limnohabitans* | NA | 0.038 | 0.005 | 0.055 | 0.111 | 0.071 | 0.441 | 0.055 | 0.037 | 0.150 | 0.062 | 0.084 | 0.059 | 0.242 | 0.040 | 0.494 | 0.056 | 0.087 | 0.060 | 0.032 |
| ASV20 | Bacteria | Proteobacteria | Gammaproteobacteria | Burkholderiales | Burkholderiaceae | *Polynucleobacter* | NA | 0.031 | 0.000 | 0.045 | 0.001 | 0.059 | 0.000 | 0.047 | 0.046 | 0.000 | 0.076 | 0.003 | 0.042 | 0.000 | 0.033 | 0.000 | 0.088 | 0.001 | 0.067 | 0.002 |
| ASV21 | Bacteria | Proteobacteria | Gammaproteobacteria | Burkholderiales | Comamonadaceae | *Limnohabitans* | NA | 0.066 | 0.001 | 0.060 | 0.003 | 0.053 | 0.000 | 0.057 | 0.062 | 0.000 | 0.064 | 0.000 | 0.040 | 0.001 | 0.031 | 0.000 | 0.051 | 0.001 | 0.040 | 0.000 |
| ASV22 | Bacteria | Planctomycetota | Planctomycetes | Gemmatales | Gemmataceae | NA | NA | 0.009 | 0.170 | 0.006 | 0.138 | 0.002 | 0.003 | 0.002 | 0.003 | 0.020 | 0.003 | 0.046 | 0.009 | 0.001 | 0.006 | 0.001 | 0.001 | 0.013 | 0.001 | 0.002 |
| ASV23 | Bacteria | Proteobacteria | Gammaproteobacteria | Burkholderiales | Burkholderiaceae | *Polynucleobacter* | difficilis | 0.056 | 0.000 | 0.072 | 0.003 | 0.048 | 0.000 | 0.049 | 0.051 | 0.000 | 0.049 | 0.000 | 0.031 | 0.000 | 0.031 | 0.000 | 0.063 | 0.001 | 0.046 | 0.000 |
| ASV24 | Bacteria | Proteobacteria | Gammaproteobacteria | Burkholderiales | Comamonadaceae | *Hydrogenophaga* | palleronii | 0.001 | 0.011 | 0.002 | 0.029 | 0.018 | 0.144 | 0.005 | 0.003 | 0.009 | 0.004 | 0.022 | 0.006 | 0.097 | 0.006 | 0.055 | 0.003 | 0.047 | 0.004 | 0.007 |
| ASV26 | Bacteria | Proteobacteria | Gammaproteobacteria | Burkholderiales | Comamonadaceae | *Hydrogenophaga* | NA | 0.001 | 0.001 | 0.001 | 0.008 | 0.001 | 0.022 | 0.001 | 0.001 | 0.009 | 0.002 | 0.106 | 0.006 | 0.113 | 0.015 | 0.090 | 0.008 | 0.033 | 0.004 | 0.015 |
| ASV27 | Bacteria | Bacteroidota | Bacteroidia | Cytophagales | Spirosomaceae | *Pseudarcicella* | NA | 0.057 | 0.003 | 0.068 | 0.003 | 0.030 | 0.000 | 0.045 | 0.071 | 0.000 | 0.050 | 0.000 | 0.046 | 0.000 | 0.026 | 0.000 | 0.014 | 0.000 | 0.023 | 0.006 |
| ASV28 | Bacteria | Proteobacteria | Gammaproteobacteria | Burkholderiales | Methylophilaceae | *Methylotenera* | NA | 0.003 | 0.003 | 0.003 | 0.015 | 0.003 | 0.063 | 0.002 | 0.001 | 0.001 | 0.001 | 0.055 | 0.005 | 0.114 | 0.004 | 0.084 | 0.006 | 0.044 | 0.001 | 0.009 |
| ASV29 | Bacteria | Actinobacteriota | Actinobacteria | Frankiales | Sporichthyaceae | *Candidatus Planktophila* | NA | 0.024 | 0.004 | 0.029 | 0.007 | 0.028 | 0.000 | 0.028 | 0.040 | 0.001 | 0.075 | 0.003 | 0.050 | 0.003 | 0.043 | 0.002 | 0.058 | 0.004 | 0.051 | 0.000 |
| ASV3 | Bacteria | Cyanobacteria | Cyanobacteriia | Chloroplast | NA | NA | NA | 0.004 | 0.185 | 0.003 | 0.150 | 0.002 | 0.008 | 0.001 | 0.003 | 0.084 | 0.003 | 0.097 | 0.003 | 0.042 | 0.011 | 0.012 | 0.007 | 0.247 | 0.010 | 0.411 |
| ASV30 | Bacteria | Cyanobacteria | Vampirivibrionia | Obscuribacterales | Obscuribacteraceae | *Candidatus Obscuribacter* | NA | 0.003 | 0.049 | 0.006 | 0.064 | 0.044 | 0.014 | 0.007 | 0.007 | 0.000 | 0.010 | 0.081 | 0.024 | 0.000 | 0.027 | 0.000 | 0.007 | 0.053 | 0.025 | 0.003 |
| ASV31 | Bacteria | Proteobacteria | Gammaproteobacteria | Burkholderiales | Burkholderiaceae | *Polynucleobacter* | NA | 0.048 | 0.001 | 0.063 | 0.001 | 0.068 | 0.000 | 0.051 | 0.043 | 0.000 | 0.034 | 0.001 | 0.016 | 0.000 | 0.015 | 0.000 | 0.053 | 0.001 | 0.025 | 0.000 |
| ASV32 | Bacteria | Proteobacteria | Gammaproteobacteria | Burkholderiales | Comamonadaceae | *Caenimonas* | NA | 0.001 | 0.000 | 0.000 | 0.002 | 0.001 | 0.113 | 0.002 | 0.000 | 0.009 | 0.000 | 0.114 | 0.001 | 0.021 | 0.001 | 0.140 | 0.000 | 0.022 | 0.001 | 0.010 |
| ASV33 | Bacteria | Verrucomicrobiota | Verrucomicrobiae | Chthoniobacterales | Terrimicrobiaceae | *Terrimicrobium* | NA | 0.020 | 0.001 | 0.021 | 0.000 | 0.005 | 0.000 | 0.002 | 0.003 | 0.000 | 0.008 | 0.000 | 0.114 | 0.000 | 0.130 | 0.000 | 0.017 | 0.001 | 0.009 | 0.000 |
| ASV4 | Bacteria | Cyanobacteria | Cyanobacteriia | Chloroplast | NA | NA | NA | 0.001 | 0.127 | 0.001 | 0.154 | 0.000 | 0.008 | 0.001 | 0.001 | 0.036 | 0.001 | 0.140 | 0.002 | 0.017 | 0.010 | 0.024 | 0.007 | 0.316 | 0.015 | 0.352 |
| ASV5 | Bacteria | Proteobacteria | Gammaproteobacteria | Pseudomonadales | Moraxellaceae | *Acinetobacter* | NA | 0.000 | 0.000 | 0.000 | 0.001 | 0.000 | 0.000 | 0.000 | 0.000 | 0.225 | 0.000 | 0.053 | 0.000 | 0.034 | 0.000 | 0.001 | 0.000 | 0.001 | 0.000 | 0.017 |
| ASV6 | Bacteria | Planctomycetota | Planctomycetes | Isosphaerales | Isosphaeraceae | NA | NA | 0.015 | 0.201 | 0.003 | 0.101 | 0.002 | 0.005 | 0.002 | 0.005 | 0.189 | 0.012 | 0.087 | 0.033 | 0.032 | 0.042 | 0.015 | 0.004 | 0.061 | 0.010 | 0.101 |
| ASV8 | Bacteria | Actinobacteriota | Actinobacteria | Frankiales | Sporichthyaceae | *hgcI clade* | NA | 0.065 | 0.010 | 0.068 | 0.014 | 0.054 | 0.001 | 0.053 | 0.083 | 0.000 | 0.105 | 0.012 | 0.128 | 0.011 | 0.099 | 0.003 | 0.078 | 0.002 | 0.089 | 0.005 |
| ASV9 | Bacteria | Bacteroidota | Bacteroidia | Flavobacteriales | Flavobacteriaceae | *Flavobacterium* | NA | 0.157 | 0.013 | 0.100 | 0.012 | 0.105 | 0.001 | 0.152 | 0.119 | 0.000 | 0.019 | 0.002 | 0.002 | 0.011 | 0.001 | 0.001 | 0.041 | 0.009 | 0.022 | 0.000 |

#### 4. Supplementary Text

**Supplementary Text 1. Concentration x Time calculations summary**

Concentration x time (CT) is calculated using the method used by the water treatment plant, which follows the T10 method outlined by the US EPA in the Long-Term Enhanced Surface Water Treatment Rule.[1] The total CT for the contactor is the sum of the CTs in Cells 2, 3, and 4. Ozone is applied in Cells 2 and 3. Cell 2 is counter-current (flow is in the opposite direction of ozone gas flow) and Cell 3 is co-current (flow is in the same direction as ozone gas flow).

The CT is calculated using the ozone residuals measured at the effluent of Cells 2, 3, and 4 and the T10, which is calculated as described in **Equation S1**. All the contactor cells are 1,200 $ft^3$ (34 $m^3$) except for Cell 3, which is 2,400 $ft^3$ (68 $m^3$). Two contactors were in service for the full duration of the experiment. A baffle factor of 0.5 is used to calculate the T10 when the flow through a single contactor is less than 6.3 MGD (2.9 $\times 10^4$ $m^3$/day). When flow to a single contactor is greater than or equal to 6.3 MGD, 0.7 is used as a baffle factor.

The CT for Cell 2 is calculated using the average ozone residual, as shown in **Equation S2**. CT credits are only given for Cell 2 if the Cell 2 average ozone residual is greater than or equal to 0.1 mg/L.

The CT for Cell 3 uses the average of the Cell 2 and Cell 3 effluent ozone residuals, as shown in **Equation S3**. CT credits are only given for Cell 3 if the Cell 3 average ozone residual is greater than or equal to 0.1 mg/L.

The CT for Cell 4 uses the Cell 4 effluent ozone residual, as shown in **Equation S4**.

The applied ozone dose is calculated using the plant flow rate, the number of ozone contactors in operation, the percent ozone gas in the feed and the gas feed flow rate, as shown in **Equation S5**.

#### Supplementary Text 2. Flow cytometry gating strategy summary

A dual, fixed-gating strategy was used for the flow cytometry data analysis where FL3 (red fluorescence) was plotted versus FL1 (green fluorescence). Gates were developed through analysis of pre-testing ozone contactor influent and effluent samples. Separate gates for (A) total cell counts (TCC) and (B) intact cell counts (ICC) due to the interference from background and cell debris.

**Supplementary Text 3. Cellular ATP sample analysis summary**

Samples for cellular ATP (cATP) analysis were collected in 100 mL sterile bottles with excess sodium thiosulfate. cATP analysis was conducted following the manufacturer's instructions (LuminUltra Quench-Gone Aqueous (QGA) Method). Briefly, samples were filtered onto 0.2-micron syringe filters (50 mL for ozone influent and 100 mL for ozone effluent). cATP was extracted from the filters by passing 1 mL of the UltraLyse solution through them. The 1 mL was then transferred to 10 mL tubes containing 9 mL of UltraLute. Luminescence was measured in relative light units (RLU) by combining 100 µL of the final UltraLute solution and 100 µL of the Luminase solution in a test tube, swirling it three times, and placing it in the luminometer (PhotonMaster). Total cellular ATP (cATP) was calculated as shown in **Equation S6**.
